# Structural complexity of brain regions in mild cognitive impairment and Alzheimer’s disease

**DOI:** 10.1101/2023.04.14.23288586

**Authors:** Roni Tibon, Christopher R. Madan, Delshad Vaghari, Constantino Carlos Reyes-Aldasoro

**Affiliations:** School of Psychology, University Park, University of Nottingham, Nottingham NG7 2RD, UK; Department of Computer Science, School of Mathematics, Computer Sciences and Engineering, City, University of London, London EC1V 0HB, UK; Department of Psychology, University of Cambridge, Cambridge CB2 3EB, UK

**Keywords:** Alzheimer’s disease, Mild Cognitive Impairment, Magnetic Resonance Imaging (MRI), Fractal Dimensionality, Structure Complexity, Machine Learning

## Abstract

Early detection of Alzheimer’s disease (AD) has been a major focus of current research efforts to guide interventions at the earliest stages of the disease. Subtle changes to the brain might be observed with neuroimaging techniques, even before symptoms surface. We interrogated brain images obtained with Magnetic Resonance Imaging (MRI) from two large-scale dementia datasets (namely, ADNI and BioFIND) to establish the utility of fractal dimensionality (FD)—a relatively understudied measure that estimates the complexity of 3D structures (in this case, brain regions)—for the detection of AD. We show that FD can be used to detect group differences between patients and healthy controls, with the former showing significantly reduced complexity across multiple brain regions. Furthermore, these measures were successful when used as features for individual-based classification and were highly consistent across the two datasets. Finally, the contribution of specific brain regions to individual-based classification adhered to previous literature on the properties of the brain’s memory network. Taken together, the study offers novel and interpretable evidence for the utility of FD for the detection of AD.

## Introduction

Alzheimer’s disease (AD) is an age-related neurodegenerative disorder characterised by progressive dementia, from mild memory impairment to global cognitive dysfunction and eventually death (Goedert and Spillantini, 2006). Clinical diagnostic criteria include clinical examination and neuropsychological assessment, often informed further by biological markers including the identification of dementia and Alzheimer’s phenotype (Blennow et al., 2006), abnormalities in Magnetic Resonance Imaging (MRI) and Positron Emission Tomography (PET) neuroimaging, and biochemical changes reflected in cerebrospinal fluid (CSF), to obtain a more definite diagnostic (Dubois et al., 2007). Before the onset of AD, some individuals would experience mild cognitive changes; changes that go beyond what is expected for their age and education (i.e., outside the range for healthy cognitive ageing), but might not interfere significantly with their daily activities. The cognitive profile describing these changes was termed mild cognitive impairment (MCI)—a common early (prodromal) stage of AD, with a high probability of progression to dementia (Petersen, 2009). The diagnostic criteria for MCI rely on cognitive symptoms and performance on cognitive tests, but do not require any evidence of specific biomarkers that would indicate underlying AD pathology (Winblad et al., 2004). Although the stark cognitive impairments associated with AD might have a later onset, it had been suggested that MCI patients may already have subtle brain changes that are identifiable with neuroimaging (e.g., Cabral et al., 2015; Chincarini et al., 2011; Jack et al., 2010; Vaghari et al., 2022a). Therefore, in recent decades, substantial research efforts have been dedicated to characterising the abnormalities that can be observed in neuroimaging, in search for reliable biomarkers of MCI and AD (Reviewed, e.g., by Frisoni et al., 2010; Frizzell et al., 2022; Rathore et al., 2017).

Recently, open large-scale datasets offer additional opportunities to tackle the important issue of early diagnosis. For example, the Alzheimer’s Disease Neuroimaging Initiative (ADNI) dataset, established in 2003, is an open dataset that includes MRI T1-weighted scans from N∼500 participants (across multiple sub-cohorts) grouped into AD patients, MCI patients and healthy controls (HC; Jack et al., 2008). A more recent resource is the BioFIND dataset, which includes T1-weighted scans from N∼320 participants grouped into MCI patients and HC (Vaghari et al., 2022a). Whilst the latter BioFIND dataset was launched only recently, and for the time being only yielded two publications (Bruña et al., 2022; Vaghari et al., 2022b), the ADNI dataset was extensively studied over the last decades, yielding, according to the project’s website (https://adni.loni.usc.edu/), more than 3,000 published peer-reviewed articles. In one, relatively early but comprehensive examination, Cuingnet et al. (2011) used 10 different methods for feature extraction from structural MRIs (including voxel-based methods, cortical thickness methods, and hippocampus-based methods) in order to classify AD patients, MCI patients, and HC. The study showed better classification of AD vs. HC using whole-brain (i.e., either cortical thickness or voxel-based method) than hippocampus-based methods, and that classification accuracy drops sharply for the detection of prodromal AD (i.e., MCI vs. HC).

Despite this productive usage of ADNI data, the enthusiasm might be tempered by the problematic consequences of repeatedly analysing the same data. Namely, extensive use of a single dataset enhances the risk of increasing false positives, since researchers do not correct for the increased number of multiple comparisons across studies. It also limits the ability to generalise conclusions beyond that specific data (e.g., Madan, 2022; Tibon et al., 2022). Indeed, it was recently found that studies that used an independent dataset for validation reported much lower accuracy than studies that validated models against held-out observations obtained within the same settings (Borchert et al., 2021). For example, a classifier that was trained on ADNI data and applied to patient data collected via memory clinics, found high accuracy in the training dataset (Area under the Receiver Operating Characteristic [ROC] Curve [AUC]=0.96) but markedly reduced accuracy in the clinical settings (AUC = 0.76) for AD diagnosis (Klöppel et al., 2015). Recently, several studies were conducted to assess generalisability in unseen independent research datasets (De Carli et al., 2019; Qiu et al., 2020), demonstrating the importance of validation across datasets in identifying methodological issues relevant to the overall model performance. A thorough examination of multiple datasets is therefore important for the exploration of group-level differences and is vital for accurate classification of individual cases.

As with any multidimensional data, the selection of MRI-based features to-be-used for detection of group differences and/or classification is of crucial importance. Various types of structural features extracted from MRI images have been used in the past (e.g., Gerardin et al., 2009; Klöppel et al., 2008; Vemuri et al., 2008). In some cases, the features were defined at the level of the MRI voxel throughout the entire brain (e.g., Klöppel et al., 2008) or for specific key regions (e.g., Gerardin et al., 2009), whereas in others, preliminary steps were taken to reduce the dimensionality of the feature space using different types of feature extraction and selection methods, including smoothing, voxel-downsampling, and parcellation (e.g., Fan et al., 2008a, 2008b; Magnin et al., 2009; Vaghari et al., 2022b; Vemuri et al., 2008). Novel methods for feature extractions are constantly being developed, aiming to capture additional fine-grained aspects of brain structures.

Fractal dimensionality (FD; e.g., Madan and Kensinger, 2018, 2016) forms a feature extraction method which is relatively understudied in the context of dementias research. This method is based on an index used to describe the complexity of the cortex, originally designed to quantify the structure of fractals (details below). When used to study cortical complexity, it can be applied globally to the entire cortex, using an unparcellated cortical ribbon, or to multiple regions obtained via various parcellation schemes. To our knowledge, only a handful of studies have considered this measure in the context of MCI/AD (Reviewed by Meregalli et al., 2022; Ziukelis et al., 2022). Two studies comparing FD in MCI relative to control subjects detected global reduction in FD but no regional effects (Ma et al., 2020; Ruiz de Miras et al., 2017). Nevertheless, when Ruiz de Miras et al. (2017) grouped MCI subjects according to whether they progressed to AD within the four year study period, differences were also detected in regional white-matter. Recently, Pantoni et al. (2019) showed reduced white-matter FD in small vessel disease (SVD) patients who are also diagnosed with MCI, and that this decrease predicted worse cognitive performance. More recently, however, McDonough and Madan (2021), estimated FD in individuals at risk for dementia (based on self-reported risks, e.g., subjective memory complaints, less than a high school education, mild head trauma, family history of AD, current diagnosis of hypertension or systolic blood pressure greater than 140 mmHg, etc.), but without cognitive impairment, and did not observe an association between FD values and dementia risk.

The earliest study of FD in AD (Thompson et al., 1998) found no difference in AD patients relative to healthy controls. More recent investigations, however, were able to link AD with reduced FD across grey-mater, white-matter, global, and local measurements. More specifically, using a small subset from ADNI (with N=15 in each group), King et al., (2009) estimated FD of the cortical ribbon in seven 2D slices, showing lower FD values in patients than controls. In a follow-up study, the authors used a larger subset of ADNI (N=35 in each group) to examine 3D FD, again showing lower (3D) FD of the cortical ribbon in AD patients (King et al., 2010). Two other studies observed global reduction in FD in AD patients vs. controls, alongside specific regional effects in the medial temporal lobe and the posterior cingulate cortex, the precentral and postcentral gyri, and the temporal pole (Nicastro et al., 2020; Ruiz de Miras et al., 2017). Taken together, the majority of previous studies report some reduction in FD in MCI/AD patients vs. controls. However, these studies were conducted with relatively small samples, reducing the ability to decipher potential regional effects. In addition, analyses were restricted to a single sample thereby limiting the ability of the findings to generalise beyond specific settings. Finally, the analyses focused on group-level effects, and individual-based classification was not explored.

The aim of the current study was to establish the utility of structural complexity measures obtained from T1-weighted MRI images for the detection of dementia. First, we used FD for the detection of group differences between MCI/AD patients and HC. To this end, we obtained T1-weighted MRI images from two large-scale datasets (BioFIND and ADNI), calculated structural complexity across 5 parcellation schemes using the fractal dimensionality toolbox (Madan and Kensinger, 2016), and performed group-level statistical analyses. We predicted that FD values will be lower in MCI/AD patients vs. healthy controls. Next, to investigate the ability of this measure to facilitate potential individual diagnosis, we identified brain regions in which FD values were predictive of class attribution and designated them as selected features. These features were then used for classification of individual cases. Finally, we evaluated to which extent the results remain valid and generalise beyond a specific dataset. To this end we assessed the correspondence between measures of structural complexity in BioFIND and ADNI using feature-wise correlations at the group level, and also evaluated the performance of the classifiers (trained on ADNI) when applied to the test set (BioFIND).

## Methods

### Datasets

#### ADNI

ADNI was launched in 2003 as an open dataset, with the primary goal of assessing whether biological markers (including MRI), and clinical and neuropsychological assessments, could be combined to measure the progression of MCI and AD. The acquisition protocol and the criteria used for the inclusion of participants are fully described in the ADNI protocol (see Jack et al., 2008 for full details). In short, participants were 55-90 years old, had a study partner able to provide an independent evaluation of functioning, and spoke either English or Spanish. They were all willing and able to undergo all test procedures (including neuroimaging) and agreed to longitudinal follow ups.

Cognitive impairment in this sample was assessed using the Mini-Mental State Examination (MMSE)—a 30-point questionnaire commonly used in clinical and research settings to measure cognitive impairment (Folstein et al., 1975), and the Clinical Dementia Rating (CDR)—a global summary measure ranging between 0 and 3, designed to identify the overall severity of dementia (Morris, 1993). Throughout the entire ADNI dataset, Healthy Control (HC) participants had MMSE scores between 24 and 30 (inclusive), and a CDR of zero. They were non-depressed, non-MCI, and non-demented. MCI patients had MMSE scores between 24 and 30 (inclusive), a memory complaint, objective memory loss (as measured with the Wechsler Memory Scale Logical Memory II; Wechsler, 1945), a CDR of 0.5, and absence of significant levels of impairment in other domains. AD patients had MMSE scores between 20 and 26 (inclusive), CDR of 0.5 or 1.0, and met standard criteria for probable AD (McKhann et al., 1984).

The current sample included N=152 participants (78 females) with defaced T1-weighted structural MRI images, from the ADNI1 Baseline 3T collection. In this sample, participants’ age ranged between 55 and 89 (M=74.87, SD=6.99). The HC group included N=47 participants (29 females) aged 70-86 (M=75.06, SD=3.89), the MCI group included N=72 participants (27 females) aged 55-88 (M=75.13, SD=7.97), and the AD group included N=33 participants (22 females), aged 57-89 (M=74.03, SD=8). Independent sample t-tests confirmed no significant age difference between the groups (HC vs. MCI: *t*(117)=0.005, *p*=.96; HC vs. AD: *t*(78)=0.76, *p*=.45; MCI vs. AD: *t*(103)=0.65, *p*=.52). Nevertheless, Chi-squared tests for independence showed a significant difference in gender distribution in HC vs. MCI, *χ^2^*(1)=5.75, *p*=.016, and in MCI vs. AD, *χ^2^*(1)=6.61, *p*=.01, but not in HC vs. AD, *χ^2^*(1)=0.05, *p*=.83, resulting from greater proportion of female participants in the HC and AD groups (62% and 67%, respectively) compared to the MCI group (38%). Importantly, when the two patient groups were grouped together (as was done for most subsequent analyses, see below) the analysis showed no differences in gender distribution for patients vs. HC, *χ^2^*(1)=2.37, *p*=.12.

#### BioFIND

The BioFIND project was launched recently (Vaghari et al., 2022a) as a multi-site, multi-participant MRI and magnetoencephalography (MEG) resting-state open dataset to study dementia. Data were obtained from two sites: the MRC Cognition & Brain Sciences Unit (CBU) in Cambridge, England, and the Centre for Biomedical Technology (CTB) in Madrid, Spain. MCI diagnosis was given by a clinician based on clinical and cognitive tests, self- and informant-report, and in the absence of full dementia or obvious other causes (e.g., psychiatric).

BioFIND includes data from N=324 individuals. For the current study, we excluded data from participants with no (defaced) T1-weighted structural MRI images (N=15) and for which the FreeSurfer’s reconstruction process (detailed below) failed (N=2). Thus, the current study included data from N=307 individuals (151 females) aged 52-95 (M=71.75, SD=6.94). The HC group included N=165 participants (83 females) aged 53-95 (M=71.24, SD=7.01), and the MCI group included N=158 participants (68 females) aged 52-90 (M=72.77, SD=6.77). An independent sample t-test did not reveal a significant age difference between the groups, *t*(305)=-1.93, *p*=.054, nor did a Chi-squared test for independence showed any difference in gender distribution between the two groups, *χ^2^*(1)=0.095, *p*=.76. AD patients were not included in this dataset.

### Analysis of T1-weighted MRI data

#### Preprocessing

An overview of the approaches and methods that were used is shown in Figure 1. As shown in the figure, we used the same preprocessing pipeline for both datasets. Structural MRI data were preprocessed using FreeSurfer v.7.3.0 (https://surfer.nmr.mgh.harvard.edu), to automatically segment the volumetric datasets and parcellate the cortex from the T1-weighted images (Fischl et al., 2002). FreeSurfer’s standard pipeline was used (i.e., recon-all), with no manual edits to the surface meshes. With this pipeline, a two-dimensional cortical surface is reconstructed from a three-dimensional volume. First, the skull was stripped from the anatomical image to generate a mask that only contains the brain. Then, the interface between the white matter and grey matter for both hemispheres was estimated. This initial boundary was refined and then used as a base from which recon-all extends feelers to search for the edge of the grey matter. Once this edge was reached, datasets representing the pial surface were created, an inflated derivative of these surfaces was inflated again into a sphere, normalised to a template image that contains an average of 40 subjects, and then re-shaped into an inflated surface or a pial surface. Participants’ individual surface maps were normalised to this template in order to allow the use of an atlas for the parcellation of the cortex into anatomically distinct regions.

**Figure 1.**
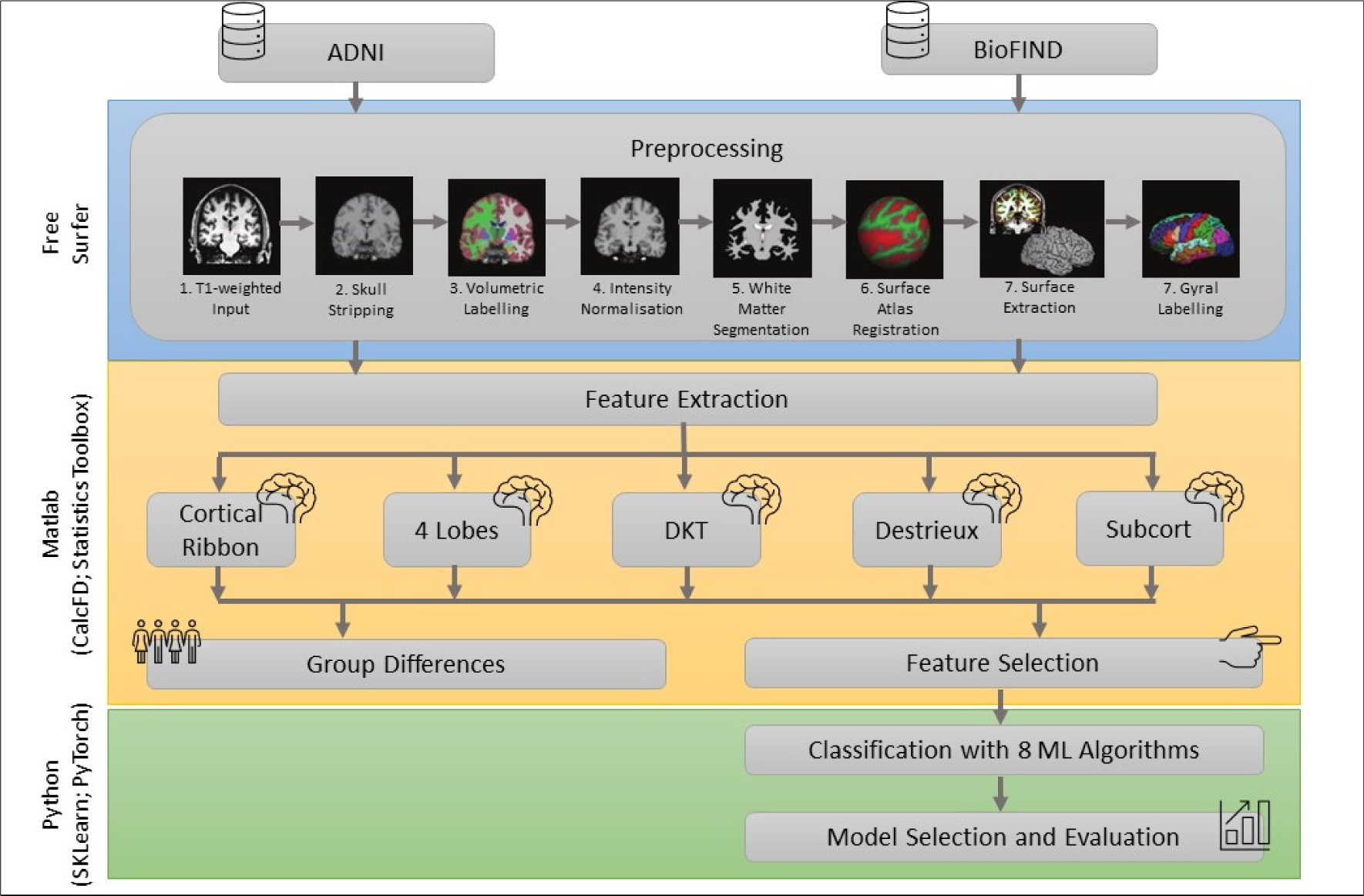
Overview of approaches and methods. Abbreviations: *ADNI*: Alzheimer’s Disease Neuroimaging Initiative; *CalcFD*: calculate fractal dimensionality; *SKLearn*: scikit-learn; *DKT*: Desikan-Killiany-Tourville; *Subcort:* subcortical; *ML*: machine learning.

#### Parcellation

Five parcellation schemes (see Figure 2) were used for feature extraction based on fractal dimensionality (FD): (1) entire cortical ribbon (one region; i.e., unparcellated); (2) each of the four lobes (four regions); (3) DKT atlas (62 regions; Klein and Tourville, 2012); (4) Destrieux et al. (2010) atlas (148 regions); and (5) subcortical structures (7 regions). The DKT and Destrieux atlases are included as standard parcellation atlases within FreeSurfer. Though some of the Destrieux regions are small, limiting the utility in measuring FD, here we chose to use it as it is well-known and readily estimable; for an alternative approach see Collantoni et al. (2020). The lobe parcellation was delineated by grouping together parcellated regions from the Destrieux atlas, as done in Madan and Kensinger (2018). Subcortical regions were defined using FreeSurfer’s automatic subcortical segmentation of the brain (aseg atlas).

**Figure 2.**
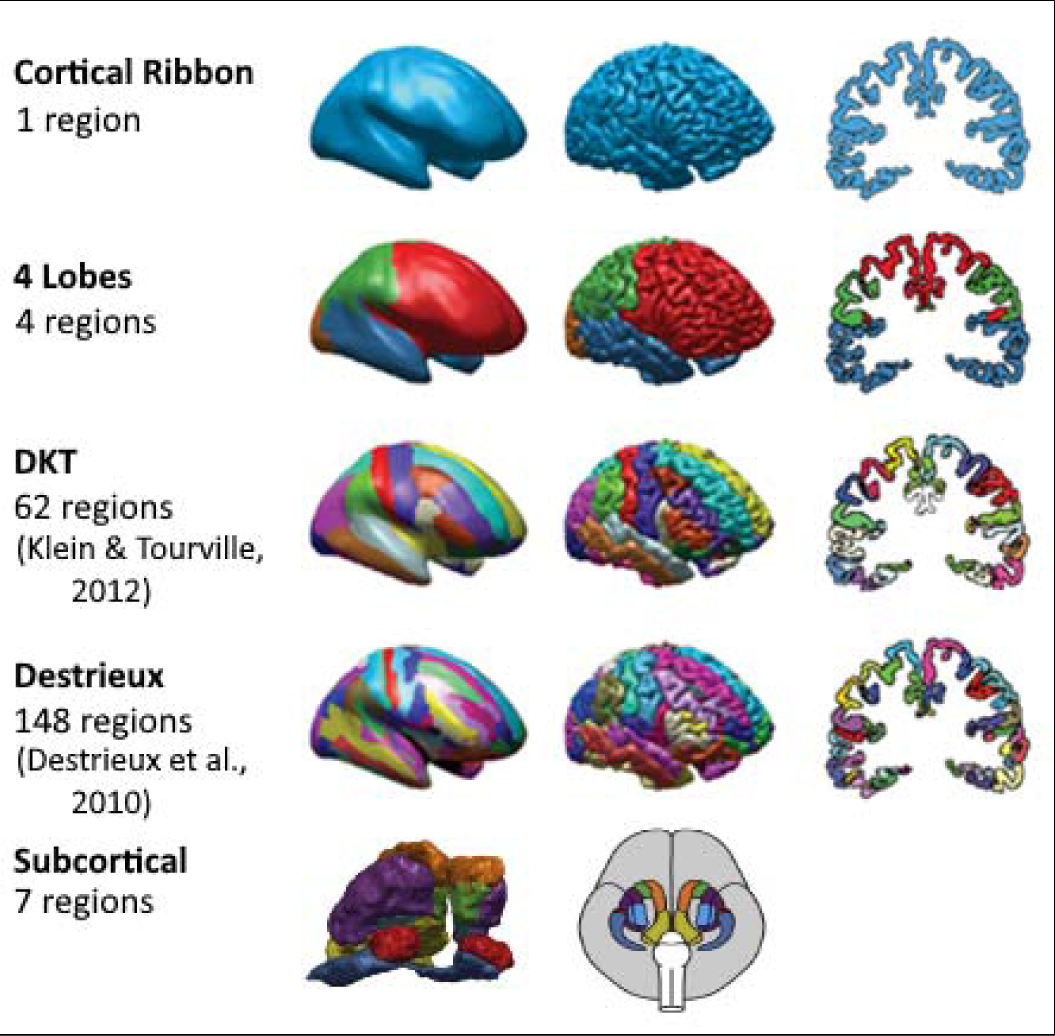
Illustration of the 5 parcellation schemes used, adapted from Madan and Kensinger (2018).

### Calculating Fractal Dimensionality (FD)

To quantify the ‘dimensionality’ or complexity of the structures (=brain regions), the Minkowski–Bouligand (see Mandelbrot, 1967) was calculated. For this calculation, the algorithm considers the 3D structure within a grid space, and the number of boxes that overlap with the edge of the structure are counted. Then, using another grid size (i.e., by changing the width of the box), the ‘box-counting algorithm’ is applied to determine the relationship between the grid size and number of counted boxes. The slope of this relationship in log-log space is the fractal dimensionality (FD) of the structure, corresponding to the equation:

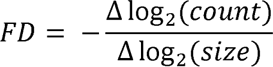

When the boxes overlapping with the edge and within the structure are both counted, the resulting slope represents the FD of the filled volume. FD was calculated using the calcFD Matlab’s toolbox (Madan and Kensinger, 2017, 2016; http://cmadan.github.io/calcFD/), which is designed to use files from the standard FreeSurfer analysis pipeline.

Using the above-described FD calculation, we extracted 10 derived datasets, one for each parcellation scheme and dataset. In each derived dataset, cases were labelled as AD (for ADNI’s datasets), MCI, and HC, according to their original classification. The number of features in each of these derived datasets was equivalent to the number of regions defined by the scheme (for example, the dataset for the DKT atlas included 64 features).

### Overall estimates of group differences

We estimated group differences in FD separately for each derived dataset using Matlab’s Statistic Toolbox (MathWorks, Inc.). For ADNI’s derived datasets, averaged FD was computed for each participant across all features and compared between groups using one-way ANOVA with group (HC, MCI, AD) as a between-subject factor. For BioFIND’s derived datasets, averaged FD was compared between groups using a two-sample T-Test.

### Feature investigation and extraction

#### General linear model and structure coefficients

To identify brain regions in which FD most reliably predicts group attribution, we computed general linear models (GLMs), separately for each derived dataset. FD values in the various parcels were used as predictors, and class as the dependent variable. Because ADNI includes two classes of patients (AD and MCI), whereas BioFIND only includes one (MCI patients), ADNI data were collapsed across groups such that for both datasets, cases were classified as either patients (PN) or healthy controls (HC). We fitted the GLMs using Matlab’s fitglm command with a binomial distribution (i.e., logistic regression, which is suitable for binary outcomes). This command outputs beta weights for the predictors, which can then be used for data description and subsequent predictions. However, when multicollinearity is present in the data, beta values are hard to interpret. Therefore, to support interpretation, we further computed structure coefficients (denoted r_s_) by correlating the values of each predictor with the predicted score. Structure coefficients can be viewed as “loadings”, indicating to what degree each variable contributes to the predictions made by the model. These coefficients can be used to guide interpretation and are particularly useful in the presence of multicollinearity (Sherry and Henson, 2005; Tibon et al., 2021; Tibon and Tsvetanov, 2022). Therefore, we used the computed structure coefficients for two main purposes, as detailed below: (1) to investigate the correspondence between the two datasets, and (2) to identify brain regions in which FD most reliably predicts group attribution.

#### Correspondence between ADNI and BioFIND

We concatenated the structure coefficients across all parcellation schemes, such that for each dataset (i.e., ADNI and BioFIND) a vector with 222 values (i.e., one r_s_ for the cortical ribbon; 4 for the 4 lobes; 62 for the DKT atlas; and so on) was generated. To assess the correspondence between the two datasets, we then correlated the r_s_ vectors generated from ADNI and BioFIND data using Pearson correlation. A high correlation would indicate that the same brain regions are predictive of group attribution in both datasets, and would therefore suggest high correspondence between them.

#### Feature extraction based on structure coefficients’ values

To decide which features should be used for machine learning (ML) classification, for each parcellation scheme, we identified features with significant r_s_ coefficients (following a Bonferroni correction across the number of parcels within the scheme). Only datasets derived from BioFIND were included in this procedure, in order to avoid any biases in the classification procedure and to allow ADNI to be used as an “unseen” dataset. That is, as described below, for classification purposes, ML algorithms were trained on BioFIND data and tested on ADNI data. Therefore, any prior selection that includes information from ADNI or information about the relations between BioFIND and ADNI, can potentially bias the results. Following this procedure, features with non-significant r_s_ values were omitted.

Whereas the brain regions comprising the subcortical parcellation scheme are unique (i.e., are not present in other parcellation schemes), other schemes include different parcellations of the same cortical regions, and therefore partially overlap. To avoid double dipping, we employed a preliminary classification procedure using logistic regression, set to select one cortical scheme to be used in subsequent analyses. Namely, for each parcellation scheme within the BioFIND dataset, we included selected features (i.e., with significant r_s_, as described above) as predictors in a logistic regression model (i.e., GLM model with binomial distribution) that classifies BioFIND’s cases into PN and HC. We then used these models, trained with BioFIND data, to classify cases in the ADNI dataset. We computed accuracy scores as the proportion of correct classifications across both groups (true positives + true negatives; shown in Table 1; results for the subcortical parcellation scheme are also shown for completeness). Note that for this preliminary step only accuracy scores were taken into account. However, additional metrics were used for the evaluation of ML algorithms, as described later on.

**Table 1.** Preliminary classification using a logistic regression algorithm for each parcellation scheme. Selected schemes are highlighted in light grey.

| Parcellation | Accuracy |
| --- | --- |
| Cortical ribbon | 0.63 |
| 4 lobes | 0.66 |
| DKT | 0.67 |
| Destrieux | 0.69 |
| Subcortical | 0.74 |

Based on these accuracy scores, we decided to use features from the Destrieux parcellation scheme (30 features with significant r_s_) and from the subcortical scheme (7 features with significant r_s_) in any further ML classification. More information about these features is provided in the Results section below. Nevertheless, because group differences (for averaged FDs) were more robust when the DKT scheme was used, the analyses were repeated with 32 features with significant r_s_ extracted from the DKT derived dataset.

### Classification Algorithms

#### Training and evaluation

We used eight classification algorithms (Decision tree, K-nearest neighbour, Linear discriminant analysis, Logistic regression, Multilayer perceptron, Naïve Bayes, Random forest, and Support vector machine; described, e.g., by Bishop, 2006; Pedregosa et al., 2011) to classify participants into their respective groups (i.e., PN, HC). All algorithms were implemented with scikit-learn v0.24.1 (Buitinck et al., 2013), aside from Multilayer perceptron algorithm which was implemented with PyTorch v1.8.0 (Paszke et al., 2019).

We designated BioFIND as the training set, due to the greater number of observations this set contains (307 vs 152 in ADNI), and better balance across groups (a 54% HC / 46% PN split in BioFIND vs. 31% HC / 69% PN split in ADNI). Therefore, each algorithm was initially trained and tuned on data from BioFIND and then tested on data from ADNI. Accordingly, BioFIND data were used for training and validation in the process of model selection. We employed an exhaustive grid search to adjust the (hyper)parameters, separately for each algorithm, with a set of relevant parameters (see Table 2). To select the best model for each algorithm (model selection), models were evaluated using a stratified 10-fold cross-validation, in which the folds are selected so that the mean response value is approximately equal in all folds. Model scoring was determined based on accuracy, calculated as the proportion of correct classifications across both groups.

**Table 2.**
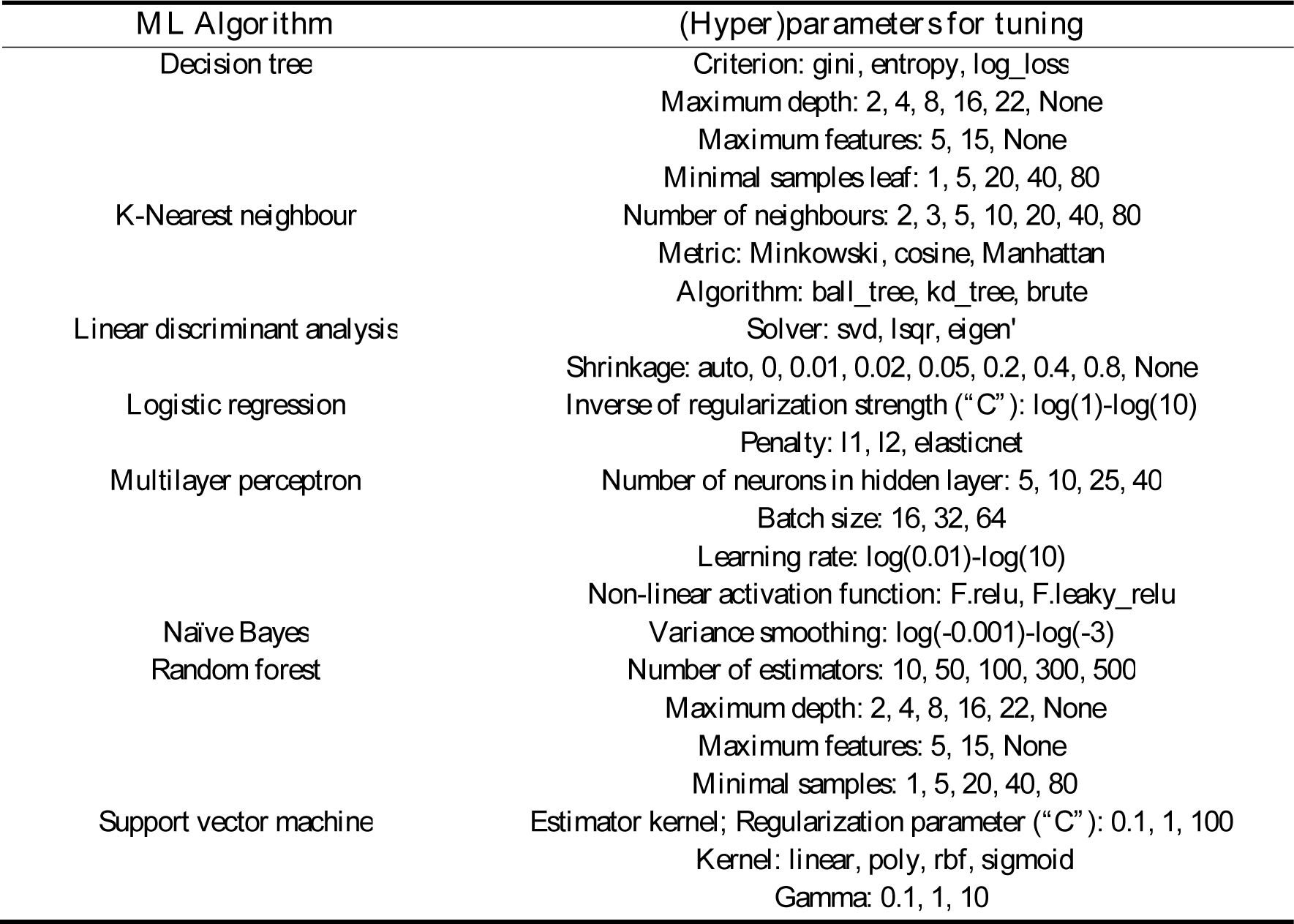
Parameters that were specified for the grid search and their selected values.

To compare how well each algorithm performs, the selected (“best”) trained model for each algorithm was then evaluated against the entire training set (BioFIND data) and, more importantly, against the unseen test set (ADNI data). The evaluation was mainly based on accuracy and AUC, that is, the two-dimensional area underneath the receiver operating characteristic (ROC) curve which shows the performance of the classification model at all classification thresholds. Nevertheless, the following additional metrics were also considered: (1) Recall, defined as the number of true positives (TP) divided by the total number of TP and false negatives (FN), i.e., 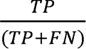; (2) Precision, defined as the number of TP divided by the total number of TP and false positives (FP), i.e., 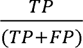; (3) F1 score, i.e., the harmonic mean of precision and recall, i.e., 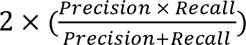, which takes both metrics into account, giving them equal weights; and (4) balanced accuracy score, defined as the average of recall obtained on each class. These measures were calculated separately for each group. In addition, confusion matrices, summarising the performance of each algorithm, and ROC curves were plotted and evaluated.

#### Further investigation

As further detailed in the Results section, based on performance, Random Forest (RF) was identified as the most successful algorithm and was therefore investigated further. This algorithm can be used to evaluate the importance of features in a classification task (e.g., Murphy, 2022), and so two methods, commonly used for this purpose, were employed here. The first is based on impurity decrease (also referred to as “gini importance”). With this method, importances are computed as the mean and standard deviation of accumulation of the impurity decrease within each tree. However, impurity-based feature importances can be misleading for high cardinality features (that is, features that can contain many unique values, as in the current case). The second common method—permutation feature importance—addresses this issue. With this technique, importance is defined as the decrease in a model score when a single feature value is randomly shuffled. This shuffling breaks the relationship between the feature and the target for the selected feature. Therefore, any drop in the model score is indicative to what extent the model depends on the feature that was shuffled. One advantage of this technique is that it does not depend on specific assumptions (i.e., model agnostic). Additionally, it can be calculated many times with different permutations of the feature thereby allowing more reliable estimates.

### Data and code availability

Raw ADNI data is available via ADNI’s project: https://adni.loni.usc.edu/, and raw BioFIND data is available via DPUK’s portal: https://www.dementiasplatform.uk/. Preprocessed (derived) data used for group analyses, ML classification, trained models, and all code used throughout the study, are available through: https://github.com/ronitibon/FDAD.

## Results

### Overall estimates of group differences

t/F-statistics, p-values, and Bonferroni corrected p-values (across all parcellation schemes) are shown in Table 3. Boxplots depicting averaged FDs for each derived dataset are shown in Figure 3. As expected, an independent sample t-test, held separately for each BioFIND’s derived dataset, revealed lower averaged FD values for MCI patients vs. HC. Similarly, for ADNI’s derived datasets, one-way ANOVA revealed a linear trend whereby averaged FD values for AD patients < MCI patients < HC.

**Figure 3.**
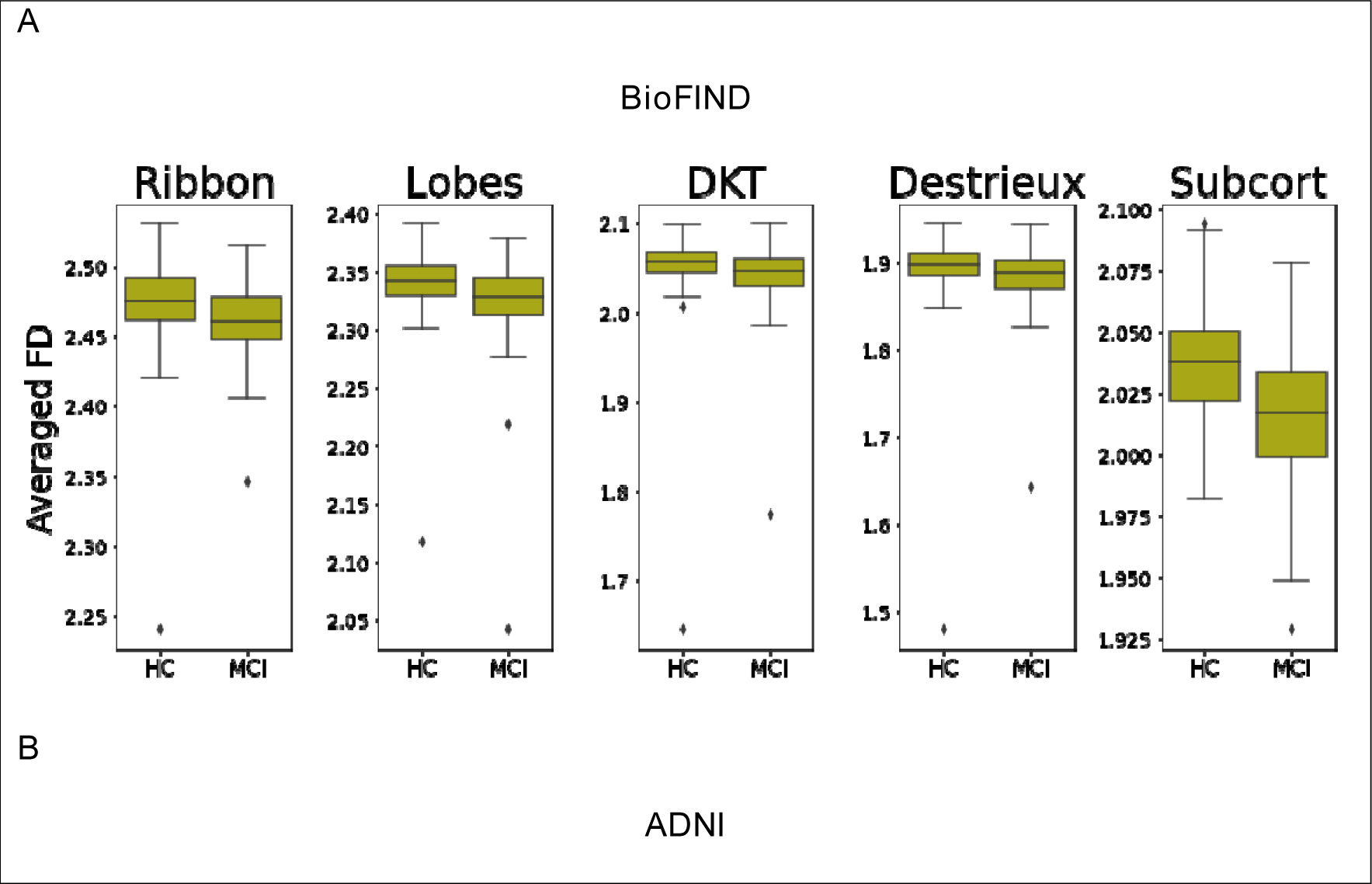
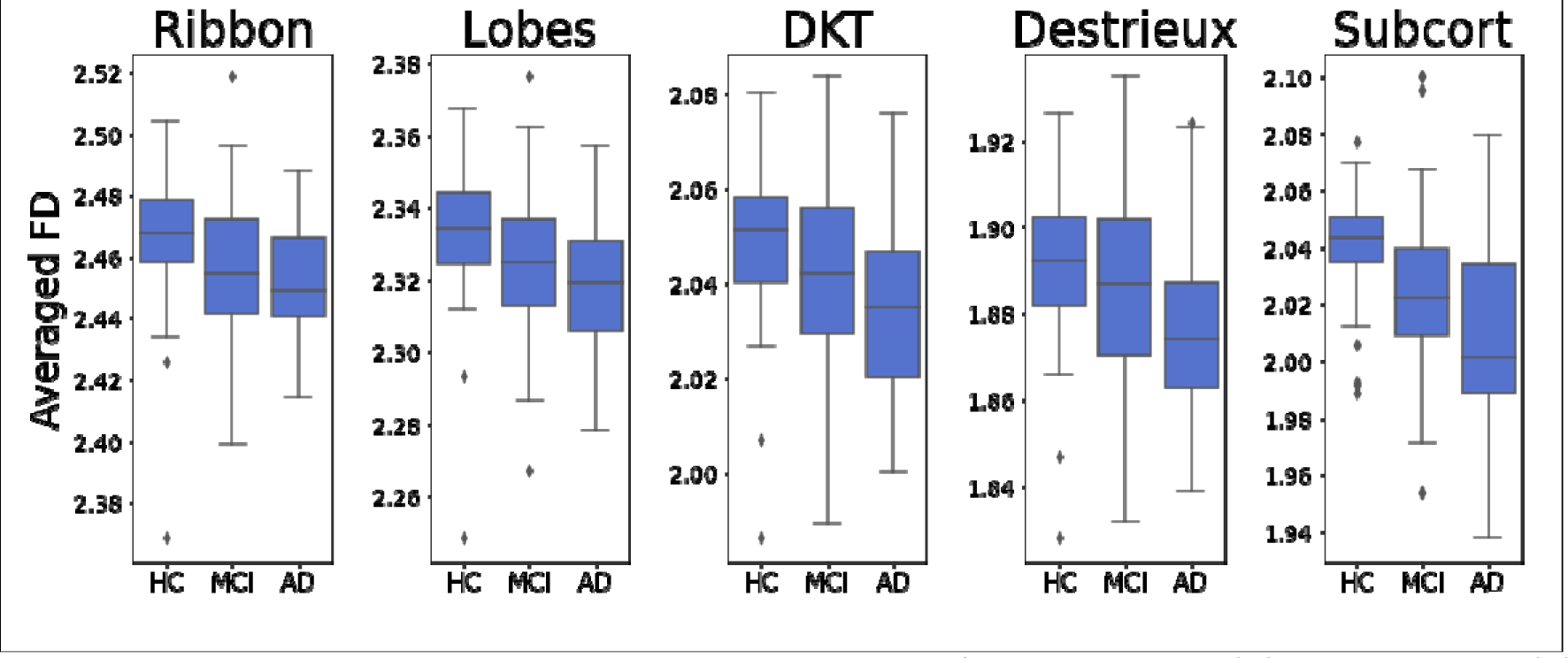
Averaged FD values across each parcellation scheme for BioFIND data (A) and ADNI data (B). HC=Healthy controls; MCI=Mild cognitive impairment; AD=Alzheimer disease.

**Table 3.** Estimates of group differences.

| Parcellation | Statistic | P-value | Corrected p-value |
| --- | --- | --- | --- |
| *BioFIND* | *t*(305) |  |  |
| Cortical ribbon | 4.69 | <.001 | <.001** |
| 4 lobes | 4.38 | <.001 | <.001** |
| DKT | 3.04 | =.003 | =.015* |
| Destrieux | 3.08 | =.002 | =.01* |
| Subcortical | 7.81 | <.001 | <.001** |
| *ADNI* | *F*(2,149) |  |  |
| Cortical ribbon | 5.31 | =.006 | =.03* |
| 4 lobes | 5.05 | =.008 | =.04* |
| DKT | 5.64 | =.004 | =.02* |
| Destrieux | 4.77 | =.01 | =.05 |
| Subcortical | 13.46 | <.001 | <.001** |

For Bonferroni corrected p-values: \*\* $p < .01$ ; \* $p < .05$

As shown in Figure 3, each dataset included a few outliers. To confirm that these outliers did not drive the results, we removed them by excluding cases with averaged FD of more than three scaled median absolute deviation (MAD) from the median of each group and repeated the analyses again. The outcomes of these additional analyses (included in Supplementary Analysis 1) showed that the removal of these outliers had very little influence and did not change any of the key results. Therefore, these cases were included in all subsequent analyses.

### Feature investigation and extraction

#### General linear model and structure coefficients

To visualise the contribution of different brain regions to the prediction of class attribution (Figure 4), we replaced values in a 3D brain volume of an exemplar participant (‘Bert’) with r_s_ values obtained from BioFIND data, and overlayed on a structural brainmask MRI image of that participant. This was done for the Destrieux and subcortical parcellation schemes, subsequently used for ML classification (see below).

**Figure 4.**
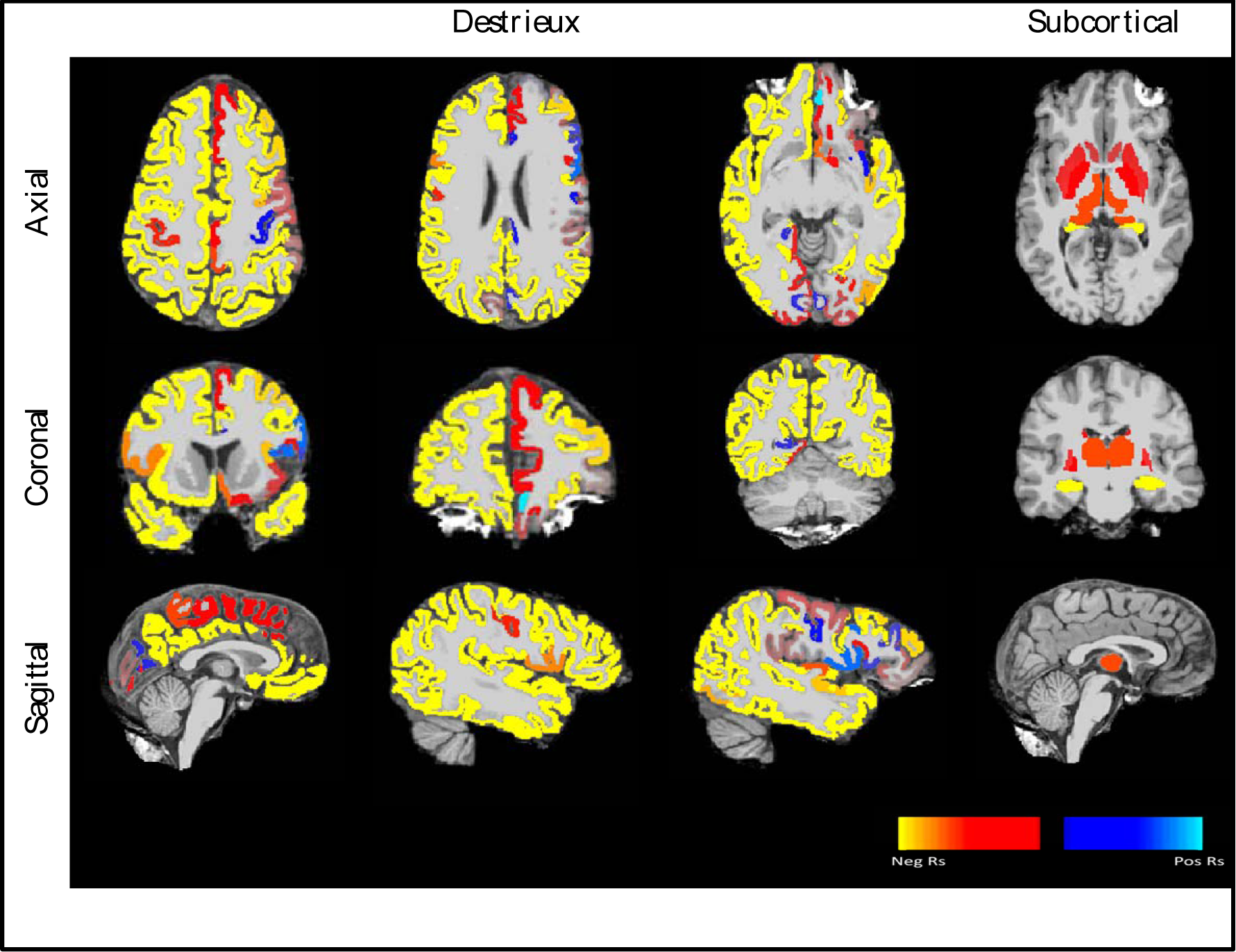
r_s_ values for the Destrieux (three left columns) and subcortical (right cloumn) parcellation schemes, overlayed on a template MRI and presented from an axial (top), coronal (middle), and sagittal (bottom) views. Warm colours indicate negative values whereas cold colours indicate positive values. The greater the absolute value is, the better the ability of the region to accurately classify individuals into their respective groups. Note that due to the labelling of the groups (i.e., in the data matrix individuals in the control group were labelled as “0” and individuals in the MCI group were labelled as “1”), negative values indicate greater structural complexity in healthy controls vs. MCI patients and vice versa. The values were adjusted to aid visualisation and therefore the exact numbers are arbitrary and not shown here.

As shown in Figure 4, negative r_s_ values were observed in most brain regions within these selected parcellation schemes, adhering, as expected, to lower FD values for MCI patients compared to HC. Within the Destrieux parcellation scheme, some positive values were also observed (for example, in the right central sulcus, in the right opercular and triangular parts of the inferior frontal gyrus, and in the right cuneus). However, as outlined below, these values were not significant and therefore were not included in subsequent ML classification. Otherwise, negative values were observed throughout the cortex, although the effect was somewhat left lateralised, with stronger (i.e., more negative) values observed in the left hemisphere. Within the subcortical parcellation, all r_s_ values were negative, and the strongest (i.e., most negative) value was observed in the hippocampi followed by the amygdalae. This scheme was implemented bilaterally, and therefore lateralisation patterns cannot be inferred. Note that due to the arbitrary scale, direct comparisons can only be made between different regions within the same parcellation scheme, and not across schemes.

#### Correspondence between ADNI and BioFIND

The relations between r_s_ obtained from BioFIND and ADNI are shown in Figure 5. As shown in the figure, and confirmed by a Pearson correlation test, the datasets were highly correlated, r=.72, *p*<.001. Nevertheless, one-sample Kolmogorov-Smirnov test ran separately for each dataset, indicated that the data are not normally distributed (for both BioFIND and ADNI: h=1, *p*<.001), rendering the use of a Pearson coefficient suboptimal. Therefore, we repeated the analysis using a Spearman coefficient instead. This analysis also revealed a highly significant (though more modest) correlation between the datasets, rho=.49, *p*<.001. Taken together, these results suggest that similar brain regions can be used to predict class attribution across the two datasets based on FD values.

**Figure 5.**
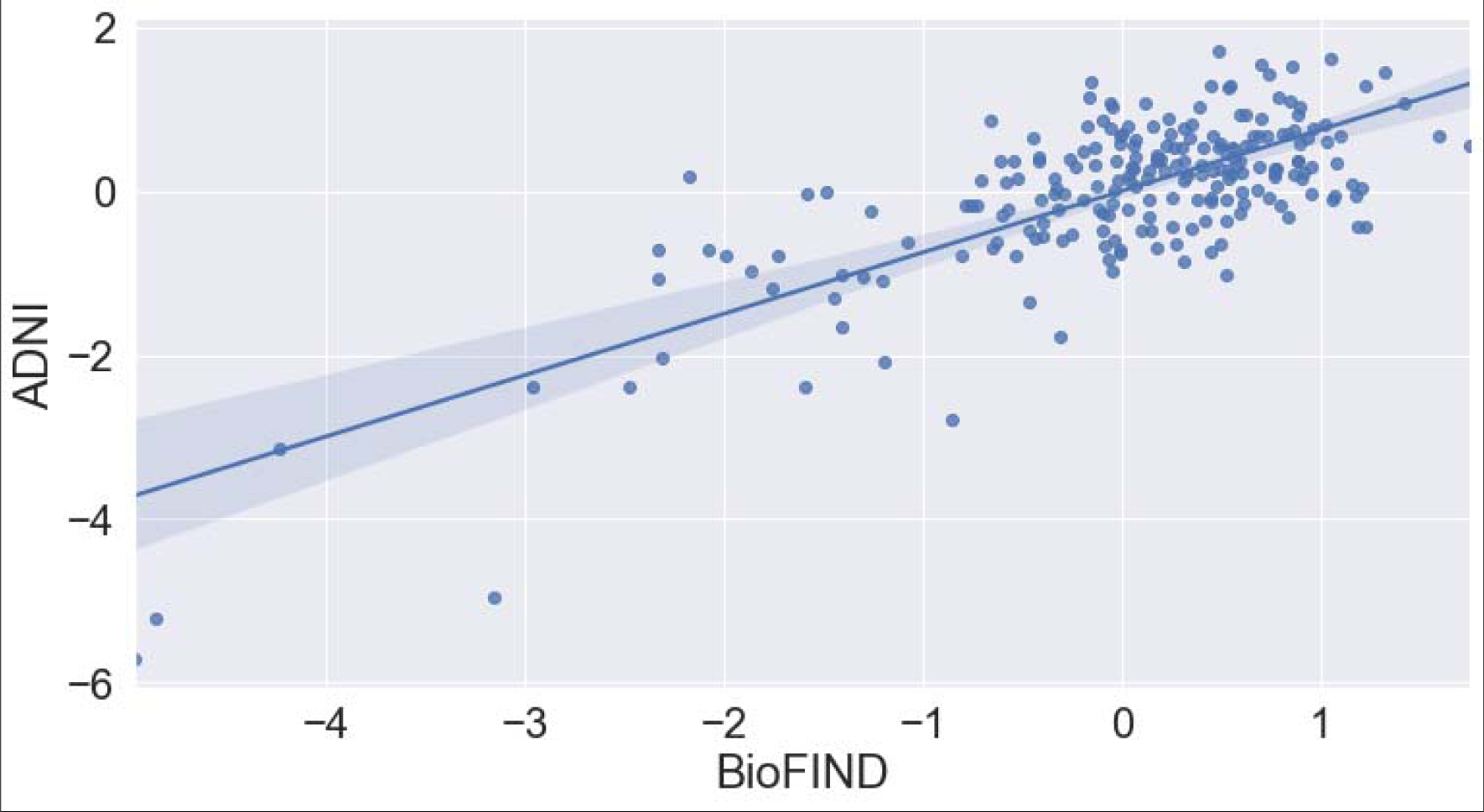
Correlation between z-scored r_s_ values in BioFIND (x-axis) and ADNI (y-axis). Each blue mark represents one brain region (i.e., one of 222 r_s_ values). Solid blue line represents the linear fit for the data, whereas blue shades indicate the confidence boundaries.

### Feature extraction based on structure coefficients’ values

A list of brain regions used for ML classification based on the procedure described in the methods section above, is shown in Table 4. Altogether, we calculated 30 features, consisting of 23 FD values obtained via the Destrieux parcellation scheme, and 7 FD values from the subcortical parcellation scheme. Thus, for each of the two datasets, a matrix with 31 columns was created: 30 features and an additional column for the target variable (class), labelled as either 0 (HC) or 1 (PN). As described above, BioFIND data included 307 cases (142 HC and 165 PN), whereas ADNI data included 152 cases (47 HC and 105 PN). These data, summarised in Figure 6, were used for ML classification.

**Figure 6.**
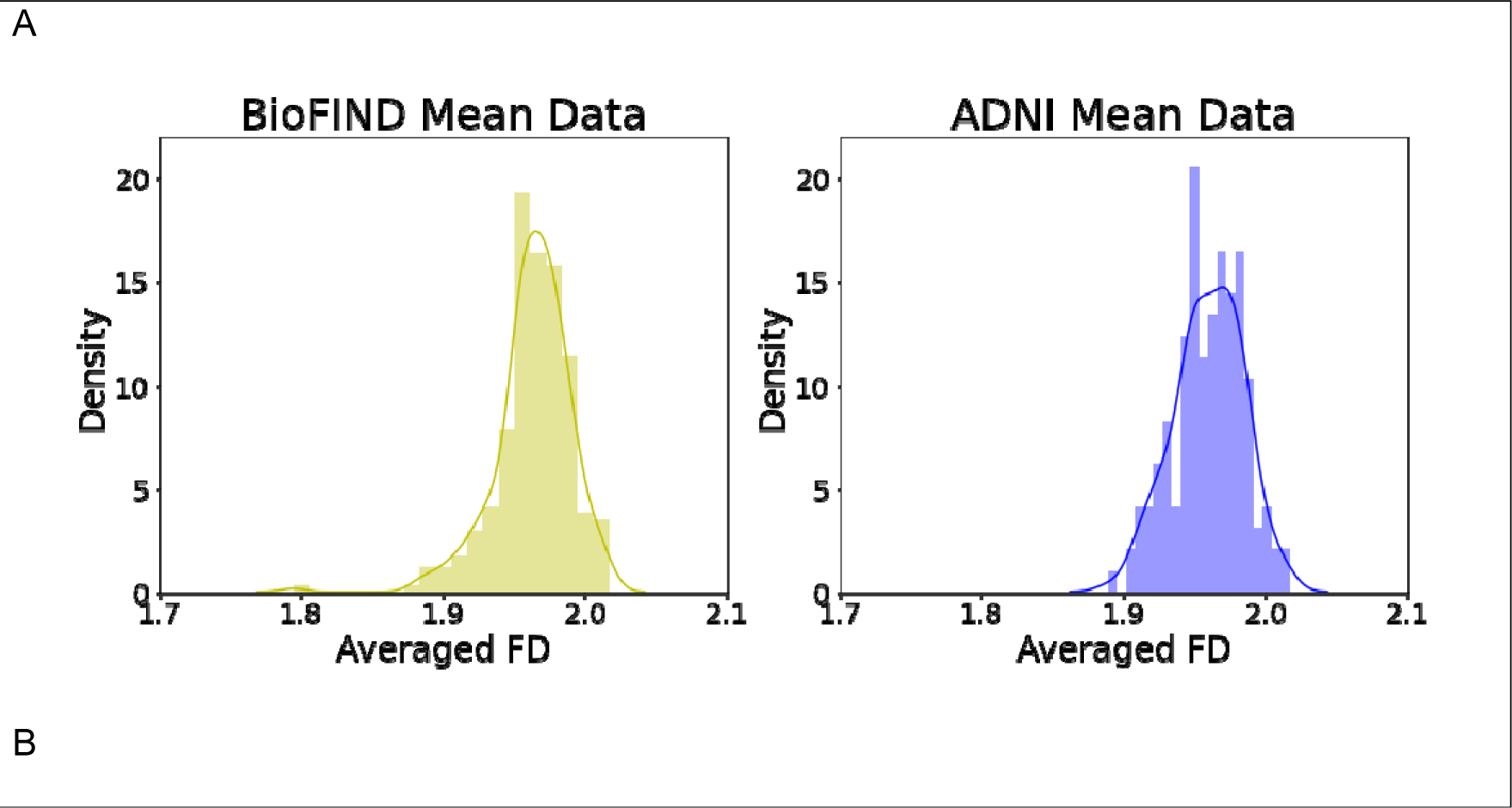
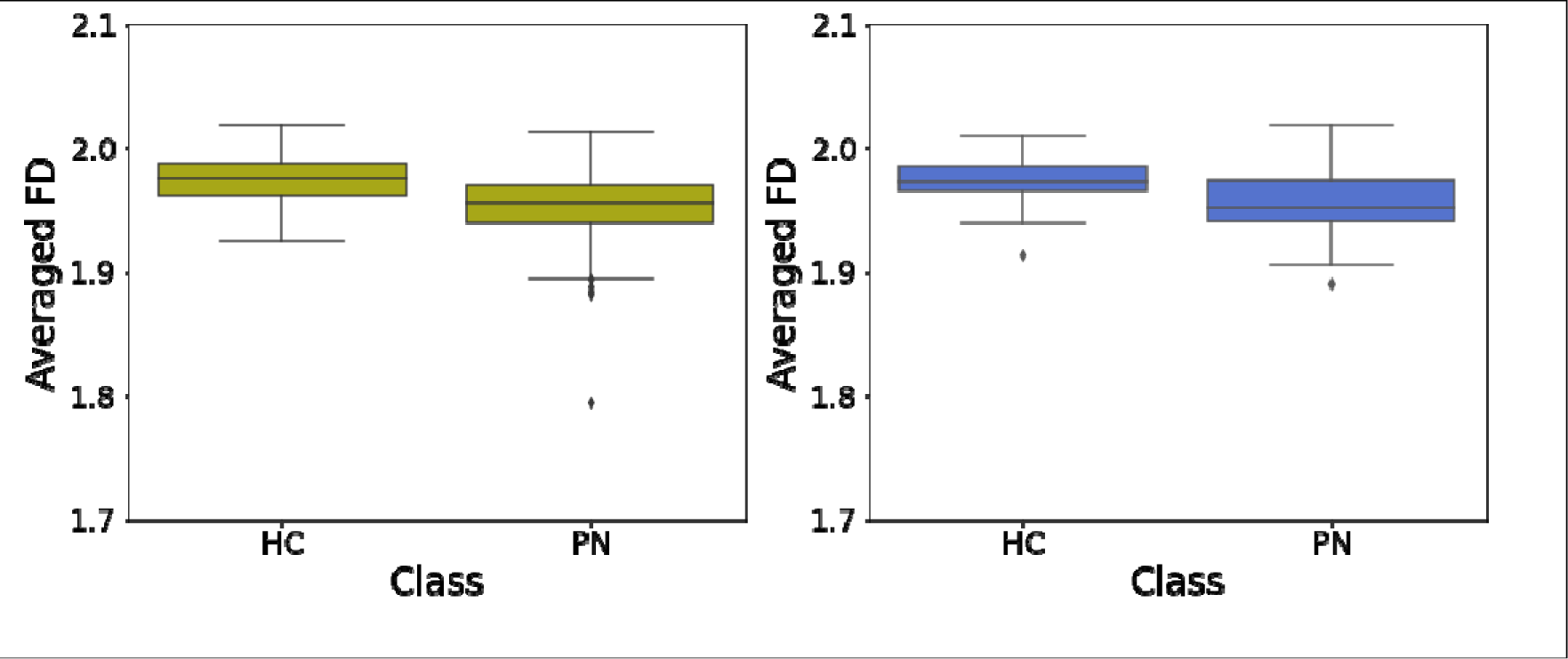
Dataset characteristics. Panel A (top) shows histograms for BioFIND (left) and ADNI (right) fractal dimensionality (FD) data, averaged across all features. Panel B (bottom) shows boxplots of averaged FD data in patients (PN) and healthy controls (HC) for each dataset.

**Table 4.**
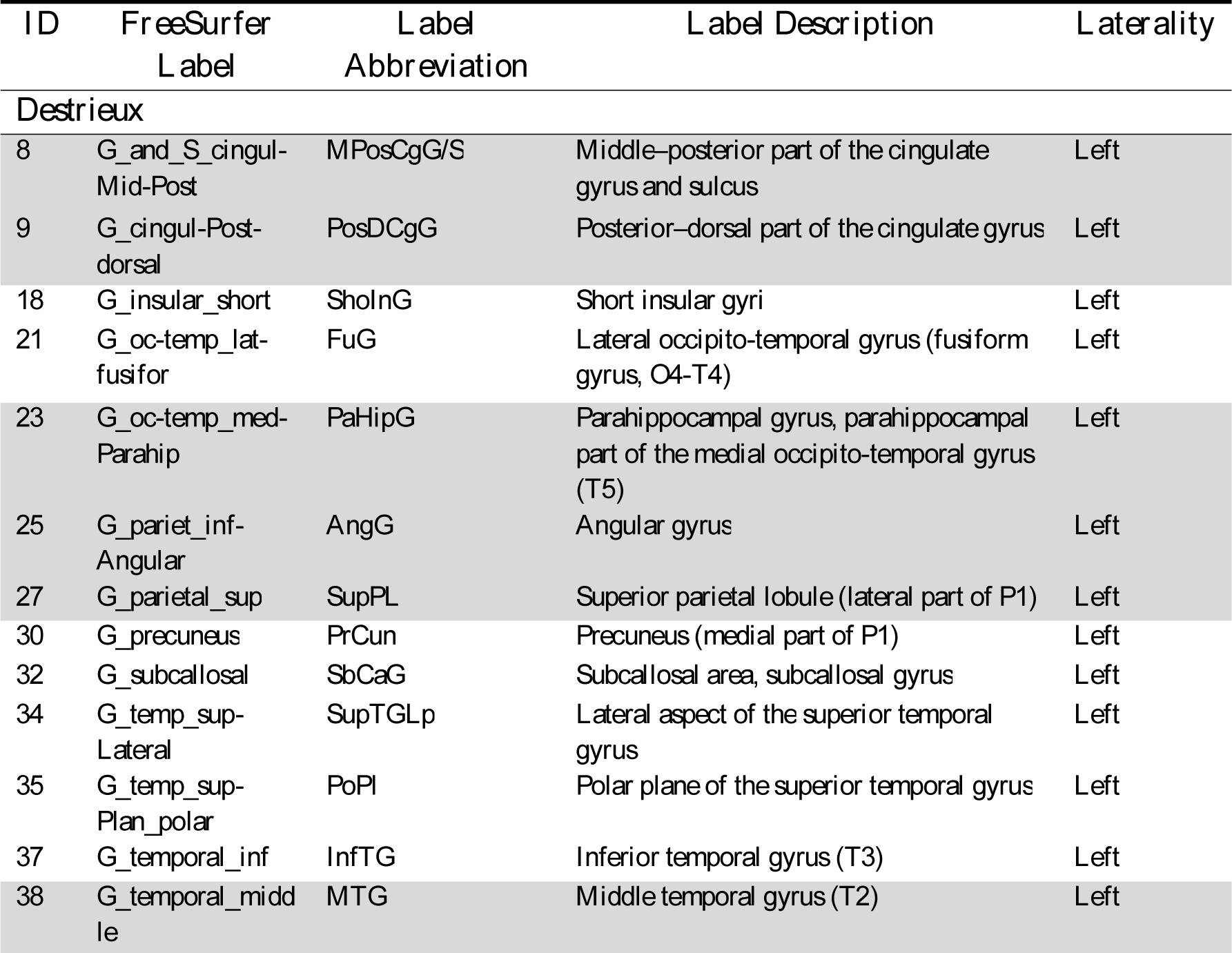
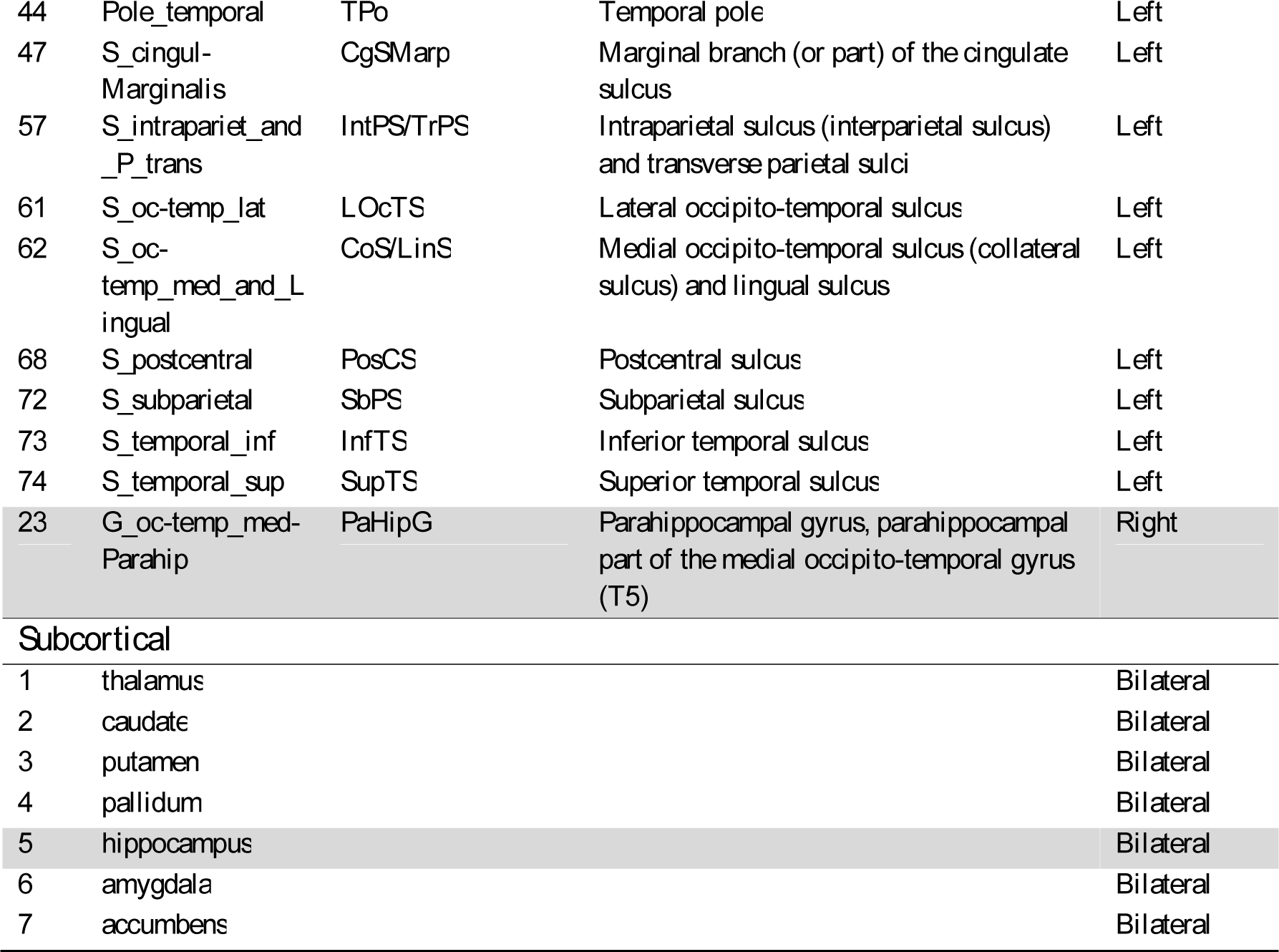
Brain regions used for ML classification. Note that (1) regions that constitute part of the brain’s memory network are highlighted with light grey shading, and (2) the left-lateralisation pattern that was observed in cortical regions (right column). These are considered in the discussion.

| ID | FreeSurfer Label | Label Abbreviation | Label Description | Laterality |
| --- | --- | --- | --- | --- |
| *Destrieux* |  |  |  |  |
| 8 | G_and_S_cingul-Mid-Post | MPosCgG/S | Middle-posterior part of the cingulate gyrus and sulcus | Left |
| 9 | G_cingul-Post-dorsal | PosDCgG | Posterior-dorsal part of the cingulate gyrus | Left |
| 18 | G_insular_short | ShoInG | Short insular gyri | Left |
| 21 | G_oc-temp_lat-fusifor | FuG | Lateral occipito-temporal gyrus (fusiform gyrus, O4-T4) | Left |
| 23 | G_oc-temp_med-Parahip | PaHipG | Parahippocampal gyrus, parahippocampal part of the medial occipito-temporal gyrus (T5) | Left |
| 25 | G_pariet_inf-Angular | AngG | Angular gyrus | Left |
| 27 | G_parietal_sup | SupPL | Superior parietal lobule (lateral part of P1) | Left |
| 30 | G_precuneus | PrCun | Precuneus (medial part of P1) | Left |
| 32 | G_subcallosal | SbCaG | Subcallosal area, subcallosal gyrus | Left |
| 34 | G_temp_sup-Lateral | SupTGLp | Lateral aspect of the superior temporal gyrus | Left |
| 35 | G_temp_sup-Plan_polar | PoPl | Polar plane of the superior temporal gyrus | Left |
| 37 | G_temporal_inf | InfTG | Inferior temporal gyrus (T3) | Left |
| 38 | G_temporal_middle | MTG | Middle temporal gyrus (T2) | Left |
| 44 | Pole_temporal | TPo | Temporal pole | Left |
| 47 | S_cingul-Marginalis | CgSMarp | Marginal branch (or part) of the cingulate sulcus | Left |
| 57 | S_intrapariet_and_P_trans | IntPS/TrPS | Intraparietal sulcus (interparietal sulcus) and transverse parietal sulci | Left |
| 61 | S_oc-temp_lat | LOcTS | Lateral occipito-temporal sulcus | Left |
| 62 | S_oc-temp_med_and_Lingual | CoS/LinS | Medial occipito-temporal sulcus (collateral sulcus) and lingual sulcus | Left |
| 68 | S_postcentral | PosCS | Postcentral sulcus | Left |
| 72 | S_subparietal | SbPS | Subparietal sulcus | Left |
| 73 | S_temporal_inf | InfTS | Inferior temporal sulcus | Left |
| 74 | S_temporal_sup | SupTS | Superior temporal sulcus | Left |
| 23 | G_oc-temp_med-Parahip | PaHipG | Parahippocampal gyrus, parahippocampal part of the medial occipito-temporal gyrus (T5) | Right |
| *Subcortical* |  |  |  |  |
| 1 | thalamus |  |  | Bilateral |
| 2 | caudate |  |  | Bilateral |
| 3 | putamen |  |  | Bilateral |
| 4 | pallidum |  |  | Bilateral |
| 5 | hippocampus |  |  | Bilateral |
| 6 | amygdala |  |  | Bilateral |
| 7 | accumbens |  |  | Bilateral |

### Machine learning classification

As described above, ML algorithms were trained and validated on BioFIND’s data, and then tested on ADNI’s data. The results of this procedure are described in the main text below.

Additional analyses are reported in the supplementary materials. First, as mentioned in the Methods section, because group differences were more robust when the DKT scheme (instead of the Destrieux parcellation scheme) was used, the analyses were repeated with 32 features extracted from the DKT derived dataset. These results are reported in Supplementary Analysis 2. Second, our models were initially trained on a scaled version of the data. However, when re-ran on an un-scaled version, performance was slightly better. Therefore, the results reported in the main text are for the non-scaled version, and results obtained with the scaled version are reported in Supplementary Analysis 3.

#### Model selection

Figure 7 depicts cross-validation accuracy for the various algorithms, following a grid search with various combinations of (hyper)parameters (see above). Accuracy was validated across 10 folds, as outlined in the methods section. Overall, aside from the MLP algorithm, all algorithms were able to provide some accurate predictions (≥ 66%). For some algorithms, accuracy rates varied significantly as a function of the specific set of parameters (e.g., for SVM), whereas for others accuracy was more stable across models (e.g., LG). This suggests that some algorithms are more influenced by specific parameter combinations and should therefore be tuned with cautious. The highest accuracies were achieved by LDA, LG, and RF, for which the best models yielded classification accuracy greater than 73%.

**Figure 7.**
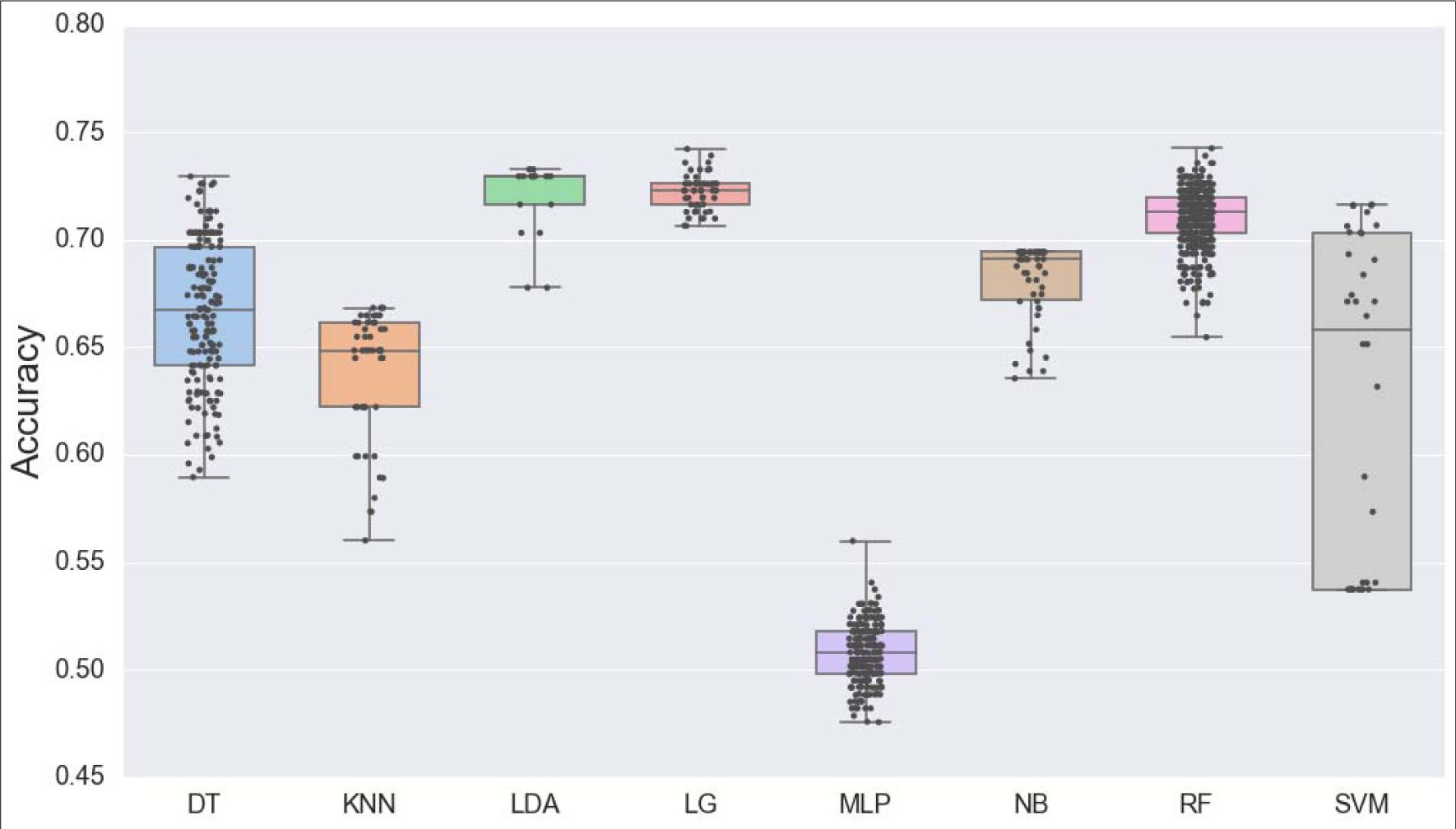
Results of the model selection process, showing above-chance performance for most algorithms. Cross-validation accuracy following a grid search with various combinations of (hyper)parameters is shown for Decision Tree (DT; blue), K-Nearest Neighbours (KNN; orange), Linear Discriminant Analysis (LDA; green), Logistic Regression (LG; red); Multilayer Perceptron (MLP; purple), Naïve Bayes (NB; brown); Random Forest (RF; pink), and Support Vector Machines (SVM; grey) algorithms.

#### Algorithm comparison

For validation of the models against the training set, the best model for each algorithm was fit on the entire training set (i.e., BioFIND data). For validation against the test set, the best model for each algorithm was fit on the unseen ADNI data. Confusion matrices for the validation against the training and testing sets, as well as the respective ROC curves, are shown in Figure 8. Additional metrics—accuracy, balanced accuracy, AUC, recall, precision, and F1 scores for the control and patient groups—were also calculated and are shown in Table 5.

**Figure 8.**
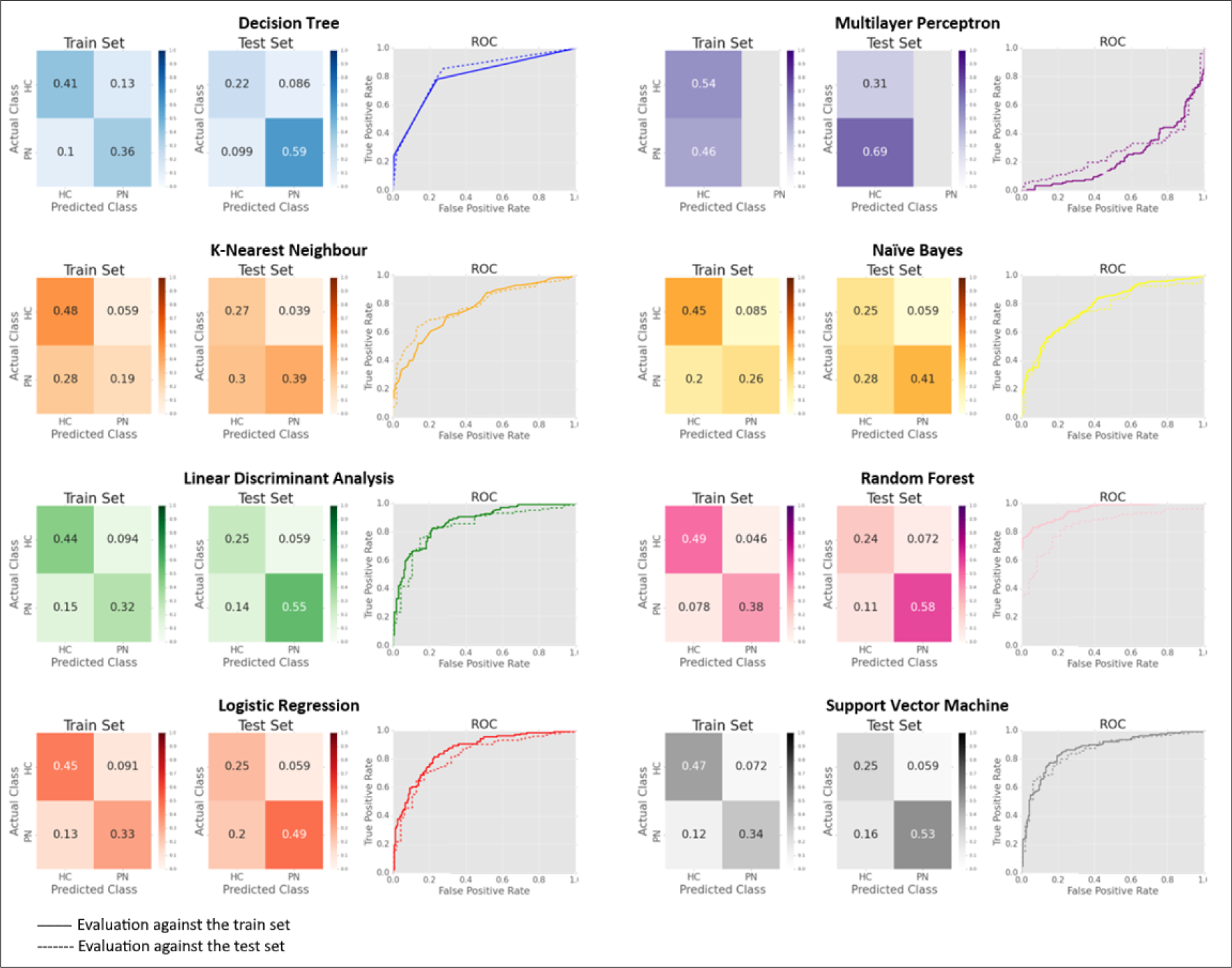
Confusion matrices and ROCs for each algorithm following the model selection procedure, showing optimal performance for the Random Forest algorithm (see text for details). Evaluation was performed against the train set (left matrix for each algorithm; solid lines in ROCs) and against the test set (right matrix, dashed lines). PN: patients, HC: healthy controls.

**Table 5.** Algorithms’ scores on various metrics. Cells are colour coded such that green colours indicate good performance and red colours indicate poor performance.

|  | **DT** | **KNN** | **LDA** | **LG** | **MLP** | **NB** | **RF** | **SVM** |
| --- | --- | --- | --- | --- | --- | --- | --- | --- |
| *Train Set (BioFIND)* |  |  |  |  |  |  |  |  |
| **Acc** | 0.77 | 0.66 | 0.76 | 0.78 | 0.46 | 0.72 | 0.88 | 0.81 |
| **bACC** | 0.77 | 0.65 | 0.75 | 0.77 | 0.5 | 0.71 | 0.87 | 0.8 |
| **AUC** | 0.8 | 0.77 | 0.87 | 0.86 | 0.24 | 0.79 | 0.95 | 0.87 |
| **RecHC** | 0.78 | 0.4 | 0.68 | 0.71 | 1 | 0.57 | 0.83 | 0.74 |
| **PreHC** | 0.74 | 0.76 | 0.77 | 0.78 | 0.46 | 0.76 | 0.89 | 0.83 |
| **F1HC** | 0.76 | 0.53 | 0.72 | 0.75 | 0.63 | 0.65 | 0.86 | 0.78 |
| **RecPN** | 0.76 | 0.89 | 0.82 | 0.83 | 0 | 0.84 | 0.92 | 0.87 |
| **PrePN** | 0.8 | 0.63 | 0.75 | 0.77 | 0 | 0.69 | 0.96 | 0.79 |
| **F1PN** | 0.78 | 0.74 | 0.79 | 0.8 | 0 | 0.76 | 0.89 | 0.83 |
| *Test Set (ADNI)* |  |  |  |  |  |  |  |  |
| **Acc** | 0.82 | 0.66 | 0.8 | 0.74 | 0.69 | 0.66 | 0.82 | 0.78 |
| **bACC** | 0.79 | 0.72 | 0.8 | 0.76 | 0.5 | 0.7 | 0.8 | 0.79 |
| **AUC** | 0.82 | 0.78 | 0.84 | 0.82 | 0.28 | 0.75 | 0.84 | 0.86 |
| **RecHC** | 0.86 | 0.56 | 0.79 | 0.7 | 1 | 0.6 | 0.84 | 0.76 |
| **PreHC** | 0.87 | 0.91 | 0.9 | 0.89 | 0.69 | 0.88 | 0.89 | 0.9 |
| **F1HC** | 0.87 | 0.69 | 0.84 | 0.79 | 0.82 | 0.71 | 0.86 | 0.82 |
| **RecPN** | 0.72 | 0.87 | 0.81 | 0.81 | 0 | 0.81 | 0.77 | 0.81 |
| **PrePN** | 0.69 | 0.47 | 0.63 | 0.55 | 0 | 0.47 | 0.68 | 0.6 |
| **F1PN** | 0.71 | 0.61 | 0.71 | 0.66 | 0 | 0.6 | 0.72 | 0.69 |

Note: Machine learning algorithms are abbreviated as DT: Decision tree; KNN: K-nearest neighbour; LDA: Linear discriminant analysis; LG: Logistic regression; MLP: Multilayer perceptron; NB: Naïve Bayes; RF: Random forest; SVM: Support vector machines; Evaluation metrics are abbreviated as Acc: accuracy; bAcc: balanced accuracy; AUC: Area under the curve; RecHC / PN: Recall of healthy controls / patients; PreHC PN: Precision of healthy controls / patients; F1HC / PN: F1 for healthy controls / patients.

As can be seen in Table 5, the RF algorithm outperformed all other algorithms (in all metrics) for the train set. This algorithm also performed well on the test set, although in this case, other algorithms (namely, DT, LDA, and SVM) performed comparably well. Therefore, the RF algorithm was explored further in order to gain additional insights into the results.

#### Further investigation

Figure 9 depicts the evaluation of feature importance using the impurity decrease technique (left) and the permutation technique (right). As can be seen, for both techniques, the hippocampus was designated as the most important feature for the model, followed by the precuneus. The inferior parietal lobe (angular gyrus) also showed relatively high values of impurity decrease, but this decrease was not significant across inter-trees variability, and was not replicated for permutation-based importances. Furthermore, the polar plane of the superior temporal gyrus, the marginal branch of the cingulate sulcus, and the postcentral sulcus, were identified as relatively important features based on permutation-based importances, but not based on impurity decrease.

**Figure 9.**
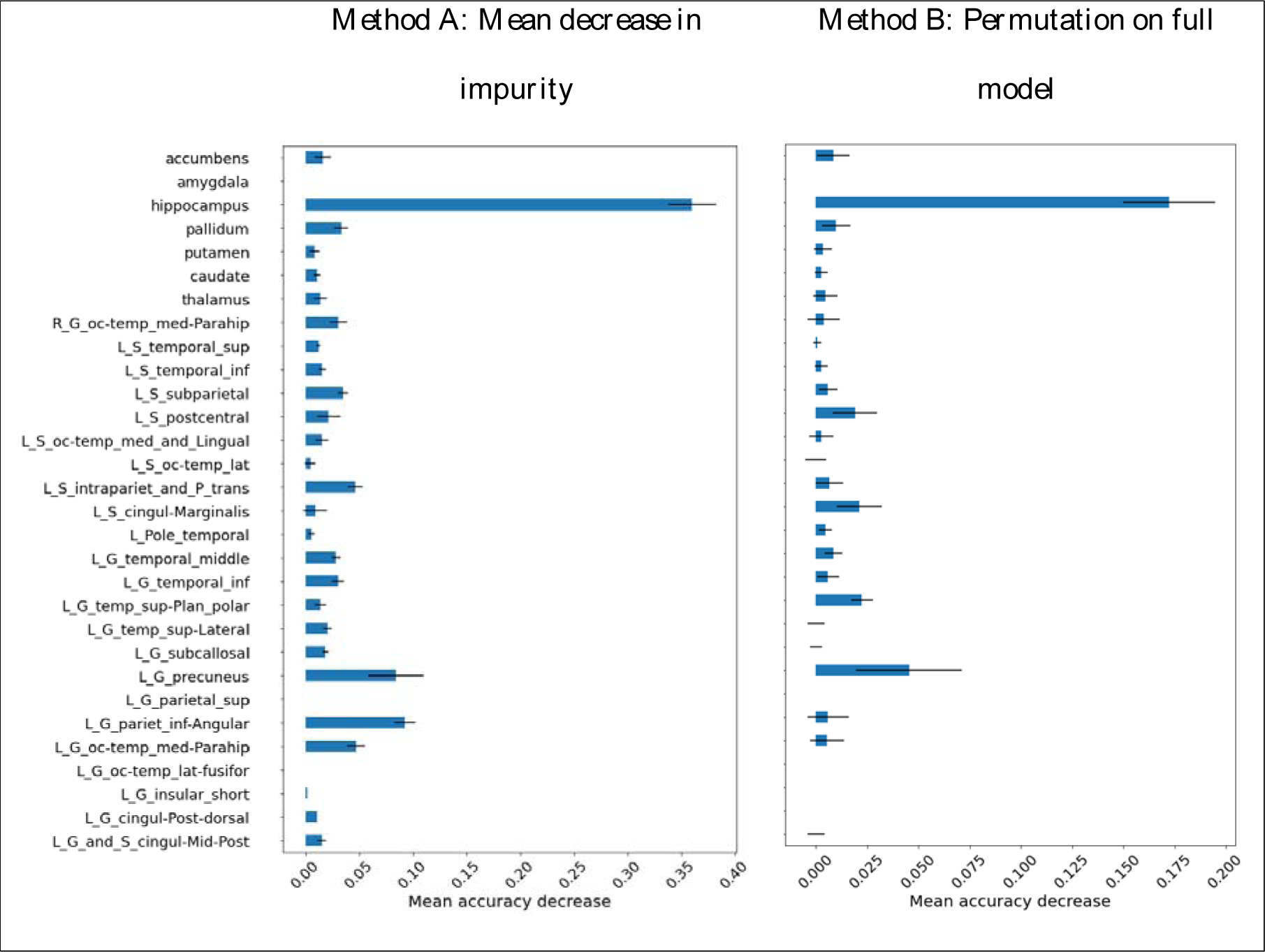
Results of feature importance evaluation using the impurity decrease (left) and permutation (right) techniques. The blue bars are the feature importances of the forest, along with their inter-trees variability represented by the black error bars.

## Discussion

The current study established FD values obtained from T1-wighted MRI images as useful measures for the detection of AD. We initially used FD to identify group differences between MCI/AD patients and healthy controls. Based on previous work (for review see Meregalli et al., 2022; Ziukelis et al., 2022) we predicted that patients would exhibit reduced structural complexity, as measured by FD values, compared to controls. This hypothesis was confirmed in the current study: for both datasets, and for all parcellation schemes, averaged FD values (across all parcels) were significantly lower in patients than in healthy controls. Moreover, in the ADNI datasets, that includes three groups (i.e., HC, MCI, and AD), we observed a linear trend whereby the averaged FD values in HC was greater than in MCI, which was in turn greater than in AD. This pattern verifies MCI as a prodromal stage of AD, during which some brain changes may already be observed, but to a lesser extent.

We then investigated whether FD can be used for AD diagnosis at the individual’s level. To this end, 8 ML algorithms were trained and evaluated on features extracted from the BioFIND dataset (using an exhaustive grid search, to select the “best model”), and then tested on parallel features extracted from ADNI. When evaluated against the training set, most models (7 out of 8) achieved accurate classification, at least to some extent (accuracy range: 0.66-0.88; AUC range: 0.77-0.95). These models were also successful when tested against the unseen test set (ADNI data; accuracy range: 0.66-0.82; AUC range: 0.75-0.86). The only algorithm that was unable to discriminate between cases in the PN and HC groups was MLP, presumably due to the small number of cases relative to the number of features. Amongst successful algorithms, random forest (RF; tuned with a maximal tree depth of 4, a maximum of 15 features, and 10 estimators) received the highest scores in all metrics when evaluated against the train set. For this algorithm, accuracy score was 0.88, AUC was 0.95, and scores in all other metrics were > 0.83. This algorithm was also successful (though to a somewhat lesser degree) when evaluated against the unseen test set (accuracy=0.82, AUC=0.84). Notably, to date, only one published study used BioFIND data for classification (Vaghari et al., 2022b). In that study, extracted features represented the mean grey matter volume across voxels within 110 cortical regions of interest, and various classification algorithms (including SVM, KNN, RF, and MLP) yielded classification accuracies ranging from 0.66-0.76; lower than those obtained here, rendering the current approach promising.

Finally, we evaluated to which extent the results remain valid and generalise beyond specific datasets. Feature-wise correlations performed at the group level indicated that similar brain regions can be used to predict class attribution across the two datasets based on their structural complexity. In addition, as mentioned above, ML classifiers trained on BioFIND data were able to correctly classify cases from the unseen ADNI dataset. Taken together, these results support good correspondence between these two datasets. This is of particular importance given that ADNI data had been widely (if not overly) used in the field, but often with limited generalisation when findings are applied to other datasets (De Carli et al., 2019; Qiu et al., 2020). The current results suggest that the measures used in the current study might be more stable across changes in settings, and therefore might be particularly useful for aiding diagnostic procedures in clinical settings.

Interestingly, some algorithms performed better when tested on unseen data than when evaluated against the training data. For example, DT’s overall accuracy scores were 0.77 for the training set but 0.82 for the test set (AUC=0.8 for train vs. 0.82 for test). Similarly, LDA’s accuracy scores were 0.76 for the train set vs. 0.8 for the test set, although for this algorithm this pattern was not observed for AUC scores, which were 0.87 vs. 0.84, respectively. Although at a first glance obtaining better performance when evaluated against unseen data might seem surprising, this pattern is reasonable given the particularities of the current datasets. Namely, the training data (extracted from BioFIND), only included MCI patients in the patient group, whereas the test data (extracted from ADNI) included both MCI and AD patients. As outlined in the introduction, MCI patients depict a specific profile of mild cognitive deficits that can evolve into the full-blown cognitive symptoms observed in AD. Similarly, mild neural changes associated with MCI can be developed into the neural pathologies that characterise AD. The current data support the notion that in some circumstances, AD can be thought of as an exaggerated version of MCI, rendering classification of AD against HC as an easier task. Therefore, the same features can be used to establish a diagnosis, but even more accurately so.

Further investigation of the contribution of specific brain regions to the observed differences between patients and controls, revealed that some regions had greater contribution to accurate classification than others (see Figure 4 and Table 4). Investigation of these regions revealed two interesting patterns. First, regions with significant contribution were mostly left lateralised. Namely, amongst the cortical regions that were selected for ML classification (based on significant structure coefficients), 22 were left-lateralised, but only one was right-lateralised. Notably, many previous studies (reviewed, e.g., by Habib et al., 2003), showed that mnemonic functions in the human brain are often lateralised to the left. This pattern adheres to the cognitive profile that characterises MCI and AD. More specifically, AD is characterised predominantly by a stark decline in episodic memory, that is, the ability to recollect events from one’s past (Dening and Sandilyan, 2015). Whilst the cognitive deficits in MCI are often more diffused, the most common subtype of MCI (∼70% of all MCI cases), termed Amnestic Mild Cognitive Impairment (aMCI), is a specific form of MCI that involves memory decline (Dubois and Albert, 2004). Although in the current datasets participants were screened for MCI and not for this specific subtype, given the high prevalence of aMCI amongst MCI cases (and for ADNI, also given that an additional inclusion criteria for the MCI group were memory complaints and objective memory loss), it is reasonable to assume that for most MCI cases an aMCI diagnosis would also be applicable. Altogether, the left-lateralisation of regions that contribute to the classification of MCI/AD patients is well aligned with the cognitive profile associated with these conditions, mainly expressed as mnemonic deficits, known to be supported by regions that reside on the left side of the brain.

A second interesting pattern concerns the contribution of specific brain regions. For several decades, regions within the medial temporal lobe—the hippocampus in particular, together with the adjacent areas including the entorhinal, perirhinal, and parahippocampal cortices—had been known as “the memory centre of the brain” and were identified as critical regions for learning and memory (e.g., Burgess et al., 2002; Eichenbaum et al., 2007; Pribram, 2012). A large body of research showed that lesions to these regions often result in severe memory deficits (e.g., Milner et al., 1998; Pribram, 2012), and that the hippocampus is especially vulnerable to damage at early stages of AD (Mu and Gage, 2011). More recently, research further identified a “core-recollection network”, that is, a network of brain regions that is applicable in various episodic memory tasks, regardless the nature of the recollected content. In addition to the hippocampus, this network includes the angular gyrus, the medial prefrontal cortex, retrosplenial/posterior cingulate cortex, precuneus, and middle temporal gyrus (King et al., 2015; Ritchey and Cooper, 2020; Thakral et al., 2017). Importantly, many of these regions (including the hippocampus, parahippocampus, angular gyrus, precuneus, cingulate cortex, and middle temporal gyrus) were identified in the current study as regions in which FD significantly contributes to classification. Moreover, investigation of the features contributing to the RF algorithm designated the hippocampus as the most important feature for the model. These, once again, reinforce the link between the measures that were used in the current study and the profile of neurocognitive deficits observed in AD/MCI.

Despite its promising results, some important limitations of the current study should be considered. First, although the datasets used in the current study are relatively ‘large-scaled’ within the domain of medical neuroimaging, in the context of ML the number of cases is highly limited. Even though performance of ML algorithms was overall promising, their accuracy falls far below any threshold that would be viable for aiding decisions in clinical settings. More data is required to improve training and to allow for more complex models, that can pick up additional subtleties in the data when making predictions. Another limitation of the current study concerns some imbalances in demographic data. Specifically, in the ADNI dataset gender was not equally distributed across groups. Given that FD values were also associated with gender, with reduced FD for females compared to males (in BioFIND: *t*(306)=6.56, *p*<.001; in ADNI: *t*(150)=2.54, *p*=.01; calculated for the averaged value across all selected features), it is possible that group differences between patients and controls are confounded by gender. However, this is unlikely to pose a concern for two reasons. First, as reported above (in the ‘participants’ section), when assessed on the concatenated data (i.e., collapsed across MCI and AD patients so that both groups are contrasted against HC), gender was evenly distributed across patients and controls. These concatenated, demographically balanced data were used for the majority of the analyses. Second, the imbalanced gender distribution in ADNI takes the form of greater proportion of females in the HC group (62%) compared to the MCI group (38%). Given the relative lower FD values in females (vs. male) and in MCI patients (vs HC), any pattern driven by a potential gender confound would attenuate the results rather than exaggerate them. In other words, the current results were obtained ***despite*** this potential confound rather than ***because*** of it; a notion that greatly alleviates this concern.

To conclude, we offer novel evidence for the utility of FD—a relatively understudied measure—in identifying group differences associated with MCI/AD. The study also provides promising direction for individual classification based on this measure. Although the translation of this work to clinical settings would require additional steps, and in particular, improving classification accuracy by additional training and tuning of the models, the correspondence that was identified here between two datasets collected in different settings points to generalisability of the current approach. Ultimately, we want to aid early diagnosis of neurodegenerative conditions in the clinical settings in which they are normally assessed. The current study entails a step in this direction.

## Data Availability

Raw ADNI data is available via ADNI project: https://adni.loni.usc.edu/, and raw BioFIND data is available via DPUK portal: https://www.dementiasplatform.uk/. Preprocessed (derived) data used for group analyses, ML classification, trained models, and all code used throughout the study, are available through: https://github.com/ronitibon/FDAD.

