## Supplementary Materials for "Structural complexity of brain regions in mild cognitive impairment and Alzheimer’s disease"

### Supplementary Analysis 1: Estimation of group differences following outliers removal

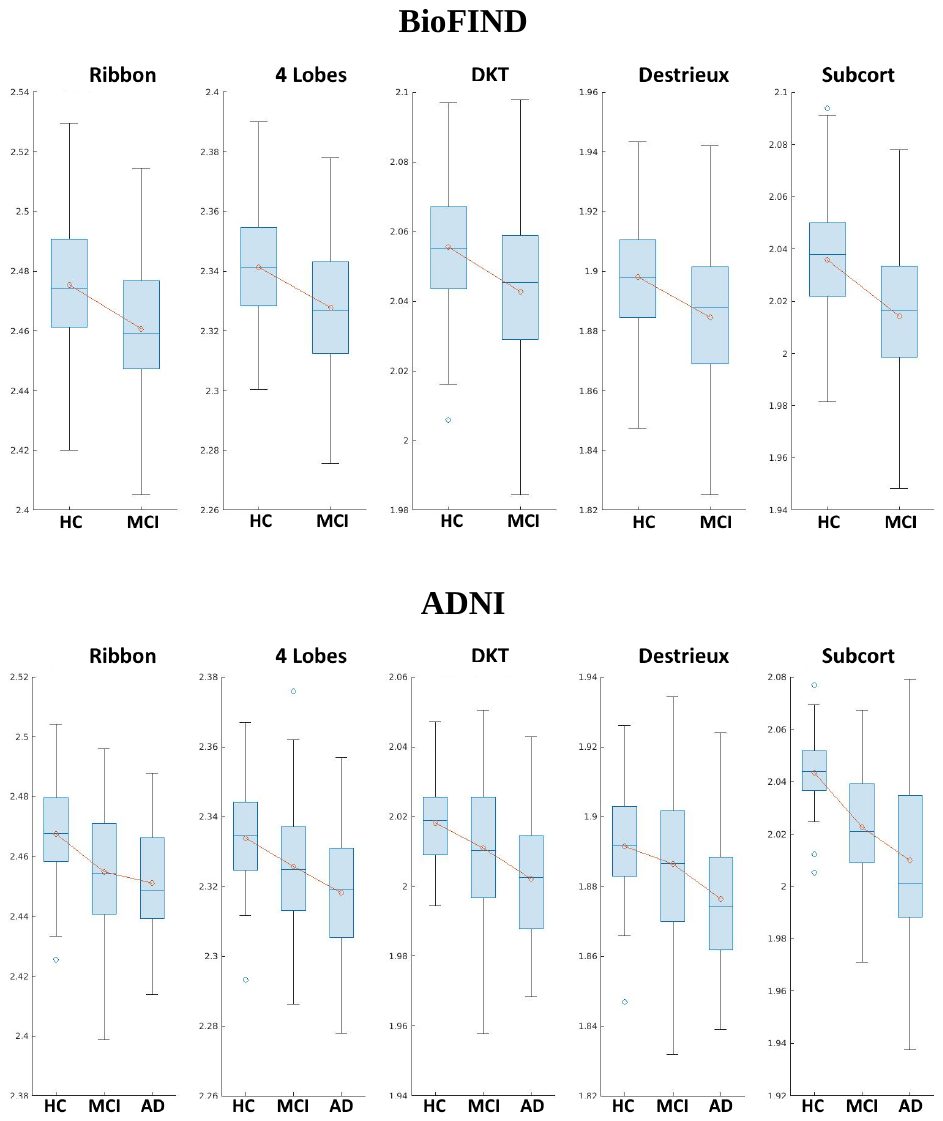

**Figure S1.** Averaged FD values for BioFIND data (top) and ADNI data (bottom) in each parcellation scheme, after the exclusion of outliers. HC=Healthy controls; MCI=Mild cognitive impairment; AD=Alzheimer disease.

**Table S1.** Estimates of group differences after removal of outliers

| **Parcellation** | **Statistic** | **P-value** | **Corrected p-value** |
| --- | --- | --- | --- |
| *BioFIND* | t(305) |  |  |
| Cortical ribbon | 6.16 | <.001 | <.001 |
| 4 lobes | 5.99 | <.001 | <.001 |
| DKT | 5.51 | <.001 | <.001 |
| Destrieux | 5.31 | <.001 | <.001 |
| Subcortical | 7.8 | <.001 | <.001 |
| *ADNI* | F(2,149) |  |  |
| Cortical ribbon | 9.61 | <.001 | <.001 |
| 4 lobes | 7.82 | .001 | .005 |
| DKT | 8.3 | <.001 | <.001 |
| Destrieux | 5.89 | .003 | .015 |
| Subcortical | 21.07 | <.001 | <.001 |

### Supplementary Analysis 2. Classification based on features extracted from DKT

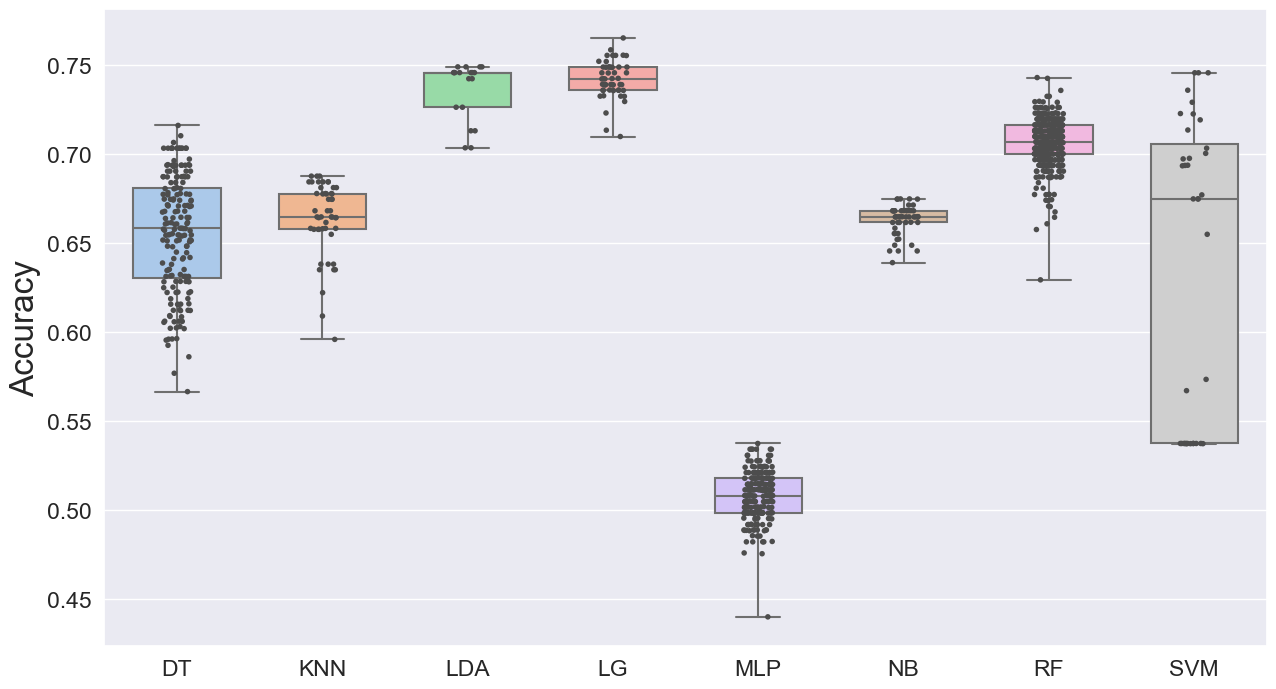

**Figure S2.1**. Results of the model selection process, with cortical features extracted from the DKT parcellation scheme.

**Table S2.** Algorithms’ scores on various metrics, with cortical features extracted from the DKT parcellation scheme.

|  | **DT** | **KNN** | **LDA** | **LG** | **MLP** | **NB** | **RF** | **SVM** |
| --- | --- | --- | --- | --- | --- | --- | --- | --- |
|  | *Train Set (BioFIND)* | | | | | | | |
| **Acc** | 0.74 | 0.71 | 0.77 | 0.8 | 0.54 | 0.68 | 0.75 | 0.79 |
| **AUC** |  |  |  |  |  |  |  |  |
| **RecHC** | 0.63 | 0.5 | 0.69 | 0.73 | 0 | 0.46 | 0.69 | 0.73 |
| **PreHC** | 0.76 | 0.81 | 0.78 | 0.82 | 0 | 0.75 | 0.75 | 0.8 |
| **F1HC** | 0.69 | 0.62 | 0.73 | 0.77 | 0 | 0.57 | 0.72 | 0.76 |
| **RecPN** | 0.82 | 0.9 | 0.84 | 0.86 | 1 | 0.87 | 0.8 | 0.84 |
| **PrePN** | 0.72 | 0.68 | 0.76 | 0.78 | 0.54 | 0.65 | 0.75 | 0.78 |
| **F1PN** | 0.77 | 0.77 | 0.8 | 0.82 | 0.7 | 0.74 | 0.77 | 0.81 |
|  | *Test Set (ADNI)* | | | | | | | |
| **Acc** | 0.78 | 0.6 | 0.71 | 0.69 | 0.31 | 0.66 | 0.77 | 0.72 |
| **AUC** |  |  |  |  |  |  |  |  |
| **RecHC** | 0.78 | 0.5 | 0.66 | 0.63 | 0 | 0.6 | 0.78 | 0.68 |
| **PreHC** | 0.88 | 0.87 | 0.9 | 0.89 | 0 | 0.86 | 0.87 | 0.9 |
| **F1HC** | 0.83 | 0.63 | 0.76 | 0.74 | 0 | 0.71 | 0.82 | 0.77 |
| **RecPN** | 0.77 | 0.83 | 0.83 | 0.83 | 1 | 0.79 | 0.74 | 0.83 |
| **PrePN** | 0.61 | 0.42 | 0.52 | 0.5 | 0.31 | 0.47 | 0.6 | 0.53 |
| **F1PN** | 0.68 | 0.56 | 0.64 | 0.62 | 0.47 | 0.59 | 0.67 | 0.65 |

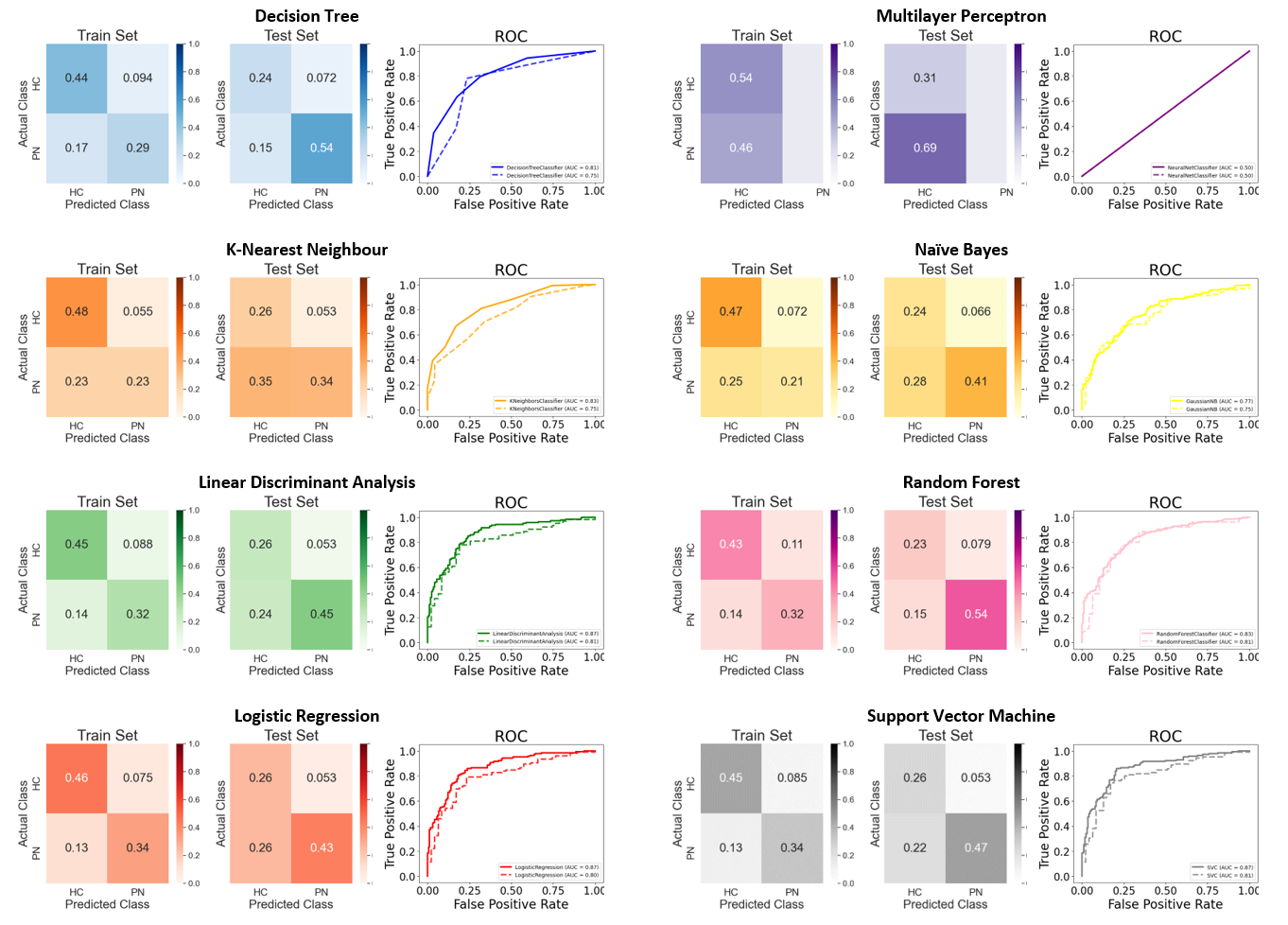

**Figure S2.2.** Confusion matrices and ROCs for each algorithm, ran on data with cortical features extracted from the DKT parcellation scheme, following the model selection procedure.

### Supplementary Analysis 3: Classification with scaled data

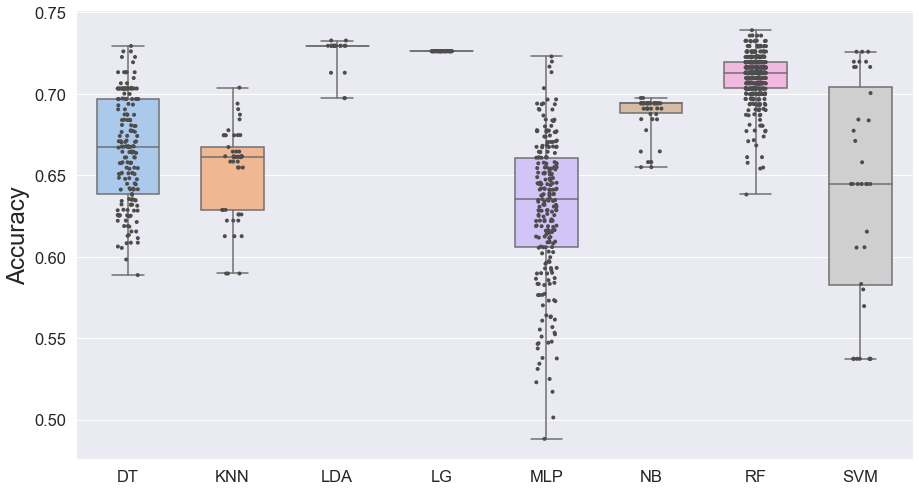

**Figure S3.1**. Results of the model selection process, with scaled (z-scored) features.

**Table S3.** Algorithms’ scores on various metrics, with scaled (z-scored) features.

|  | **DT** | **KNN** | **LDA** | **LG** | **MLP** | **NB** | **RF** | **SVM** |
| --- | --- | --- | --- | --- | --- | --- | --- | --- |
|  | *Train Set (BioFIND)* | | | | | | | |
| **Acc** | 0.74 | 0.71 | 0.77 | 0.8 | 0.54 | 0.68 | 0.75 | 0.79 |
| **AUC** | 0.81 | 0.83 | 0.87 | 0.87 | 0.5 | 0.77 | 0.83 | 0.87 |
| **RecHC** | 0.63 | 0.5 | 0.69 | 0.73 | 0 | 0.46 | 0.69 | 0.73 |
| **PreHC** | 0.76 | 0.81 | 0.78 | 0.82 | 0 | 0.75 | 0.75 | 0.8 |
| **F1HC** | 0.69 | 0.62 | 0.73 | 0.77 | 0 | 0.57 | 0.72 | 0.76 |
| **RecPN** | 0.82 | 0.9 | 0.84 | 0.86 | 1 | 0.87 | 0.8 | 0.84 |
| **PrePN** | 0.72 | 0.68 | 0.76 | 0.78 | 0.54 | 0.65 | 0.75 | 0.78 |
| **F1PN** | 0.77 | 0.77 | 0.8 | 0.82 | 0.7 | 0.74 | 0.77 | 0.81 |
|  | *Test Set (ADNI)* | | | | | | | |
| **Acc** | 0.78 | 0.6 | 0.71 | 0.69 | 0.31 | 0.66 | 0.77 | 0.72 |
| **AUC** | 0.75 | 0.75 | 0.81 | 0.8 | 0.5 | 0.75 | 0.81 | 0.81 |
| **RecHC** | 0.78 | 0.5 | 0.66 | 0.63 | 0 | 0.6 | 0.78 | 0.68 |
| **PreHC** | 0.88 | 0.87 | 0.9 | 0.89 | 0 | 0.86 | 0.87 | 0.9 |
| **F1HC** | 0.83 | 0.63 | 0.76 | 0.74 | 0 | 0.71 | 0.82 | 0.77 |
| **RecPN** | 0.77 | 0.83 | 0.83 | 0.83 | 1 | 0.79 | 0.74 | 0.83 |
| **PrePN** | 0.61 | 0.42 | 0.52 | 0.5 | 0.31 | 0.47 | 0.6 | 0.53 |
| **F1PN** | 0.68 | 0.56 | 0.64 | 0.62 | 0.47 | 0.59 | 0.67 | 0.65 |

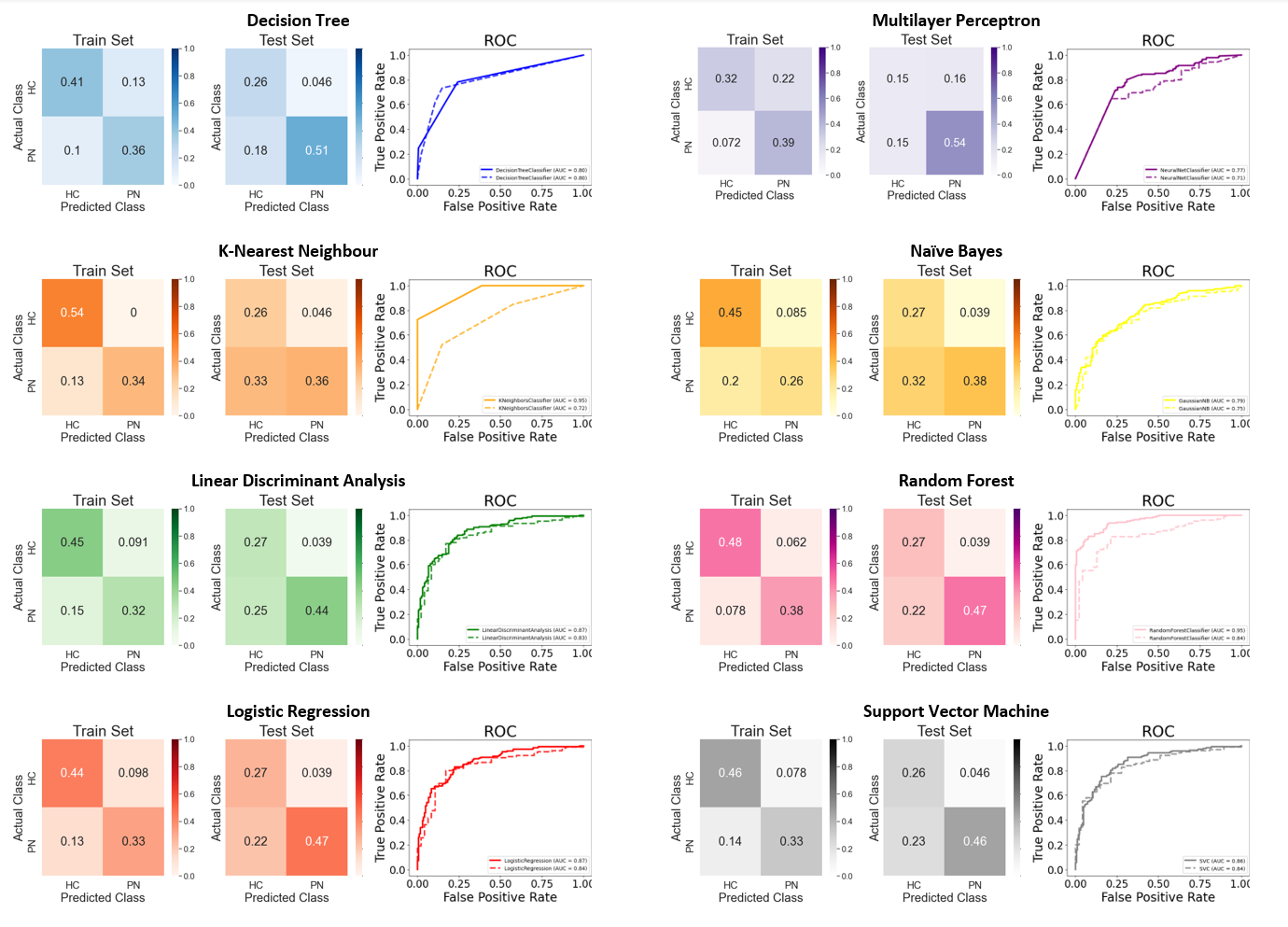

**Figure S3.2.** Confusion matrices and ROCs for each algorithm, ran on data with scaled (z-scored) features, following the model selection procedure.
